# High-throughput mass spectrometry maps the sepsis plasma proteome and differences in response

**DOI:** 10.1101/2022.08.07.22278495

**Authors:** Yuxin Mi, Katie L Burnham, Philip D Charles, Raphael Heilig, Iolanda Vendrell, Justin Whalley, Hew D Torrance, David B Antcliffe, Shaun M May, Matt J Neville, Georgina Berridge, Paula Hutton, Cyndi Goh, Jayachandran Radhakrishnan, Alexey Nesvizhskii, Fengchao Yu, GAinS Investigators, Emma E Davenport, Stuart McKechnie, Roger Davies, David JP O’Callaghan, Parind Patel, Fredrik Karpe, Anthony C Gordon, Gareth L Ackland, Charles J Hinds, Roman Fischer, Julian C Knight

## Abstract

Sepsis, the dysregulated host response to infection causing life-threatening organ dysfunction, is an unmet global health challenge. Here we apply high-throughput tandem mass spectrometry to delineate the plasma proteome for sepsis and comparator groups (non-infected critical illness, post-operative inflammation and healthy volunteers) involving 2622 samples and 4553 liquid chromatography-mass spectrometry analyses in a single batch, at 100 samples/day. We show how this scale of data can establish shared and specific proteins, pathways and co-expression modules in sepsis, and be integrated with paired leukocyte transcriptomic data (n=837 samples) using matrix decomposition. We map the landscape of the host response in sepsis including changes over time, and identify features relating to etiology, clinical phenotypes and severity. This work reveals novel subphenotypes informative for sepsis response state, disease processes and outcome, highlights potential biomarkers, pathways and processes for drug targets, and advances a systems-based precision medicine approach to sepsis.

## Introduction

Sepsis is defined as life-threatening organ dysfunction caused by a dysregulated host response to infection (Singer et al., 2016). Currently we lack effective immunomodulatory therapies to address the high mortality and global burden of this disease (van der Poll et al., 2021; Rudd et al., 2020). Incomplete knowledge of pathophysiology and failure to differentiate individual patient variation in the nature and timing of maladaptive host responses within the heterogeneous clinical syndrome of sepsis currently limit clinical trials (Marshall, 2014; van der Poll et al., 2021; Stanski and Wong, 2019). Sepsis subphenotypes informative for immune response state, outcome and therapeutic response are proposed based on clinical, laboratory and molecular stratifiers (Baghela et al., 2022; Cano-Gamez et al., 2022; Davenport et al., 2016; Kwok et al., 2022; Scicluna et al., 2017; Seymour et al., 2019; Sweeney et al., 2018; Wong et al., 2019). However, establishing the nature of the sepsis host response has been limited by incomplete knowledge of the sepsis plasma proteome. The plasma proteome reflects the function of organ systems through the secretome and tissue leakage products, offering the opportunity to identify key mediators of the sepsis response, potential therapeutic targets and biomarkers of individual variation in response.

To date there have been technological limitations to high throughput application of quantitative assays that are able to capture the high dynamic abundance range of proteins in the blood. Analysis of the sepsis plasma proteome has focused on mouse models, a small number of plasma cytokines and metabolites in patients, or mortality prediction and comparison with healthy individuals and sterile inflammation, involving relatively small numbers of patients (De Coux et al., 2015; Fjell et al., 2013; Kiehntopf et al., 2011; Langley et al., 2013; Leite et al., 2021; Miao et al., 2021; Pimienta et al., 2019; Sharma et al., 2017, 2019; Thavarajah et al., 2020; Toledo et al., 2019).

Tandem mass spectrometry (MS) provides protein measurements in an untargeted and hypothesis-free manner suitable for discovery-led characterization of the sepsis blood proteome response. Here we show how with higher-throughput automated and robust methods for sample preparation, MS-based data acquisition and data analysis it is feasible to analyze >2500 non-depleted blood plasma samples in a single batch using a liquid chromatography-MS (LC-MS) platform. We report the plasma proteome of >1000 adult sepsis patients at multiple timepoints and integrate with leukocyte transcriptomics to provide insights into the nature of the sepsis response and observed clinical heterogeneity.

## Results

### High throughput MS delineates the plasma proteome at scale for critical illness cohorts

We aimed to characterize the acute sepsis plasma proteome. To do this we analyzed patients with sepsis due to community acquired pneumonia (CAP) or fecal peritonitis (FP) admitted to the intensive care unit (ICU) and serially sampled during their admission (n=1190 patients, 1889 samples from the UK Genomic Advances in Sepsis (UK GAinS) study); and septic shock patients (n=45 patients, 154 samples from the VANISH study) (Figure 1A; Table S1; STAR Methods). Non-sepsis controls, including healthy volunteers (HV) (n=152 individuals and samples), elective surgery patients pre- and post- operation (BIONIC, X-MINS studies; n=149 patients, 351 samples) and non-infected ICU patients (MOTION, TACE and MONOGRAM studies; n=76 samples, 76 patients), were also assayed to compare sepsis with non-infectious causes of inflammation and critical illness. We defined discovery and validation cohort sets from these cases and controls (Figure 1A; Table S1; STAR Methods).

**Figure 1.**
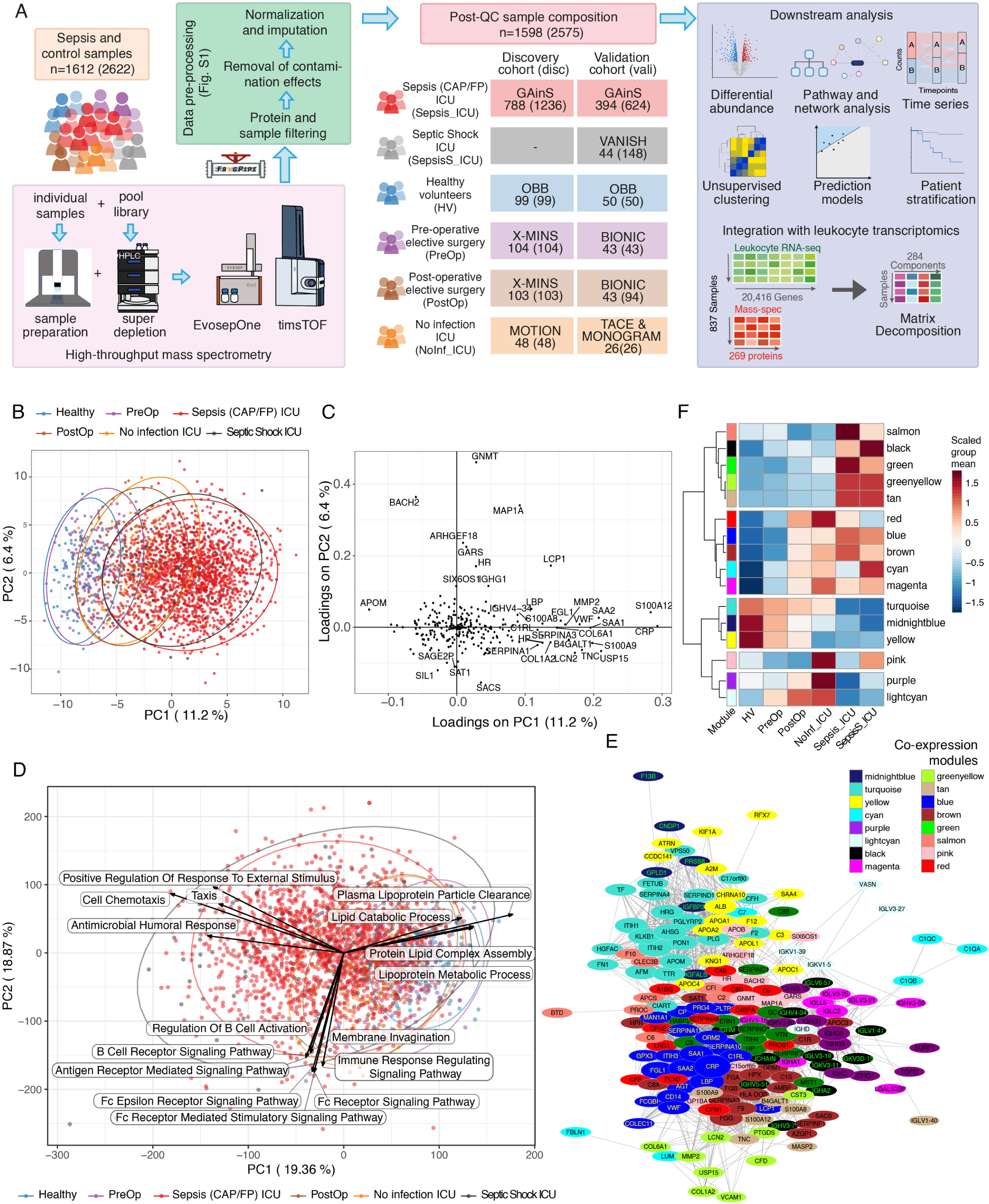
Study overview and differentiation by plasma proteome. (A) Study design, workflow and cohorts (study name, numbers of individuals assayed, and number of samples in brackets) (STAR Methods). CAP, community acquired pneumonia; FP, fecal peritonitis. (B, C) Protein abundance all samples (B) Principal component analysis (PCA) showing PC1 vs PC2 with 95% data ellipses (assuming a multivariate t-distribution) (C) protein loadings on PC1 and PC2. (D) PCA of enrichment scores matrix on all samples from Gene Set Enrichment analysis using protein abundance for single samples. Arrows, gene ontology biological processes (top 8 loadings PC1 and PC2), length scaled to loading. (E, F) Protein co-expression network from Weighted Gene Co-expression Network Analysis (E) Module network, with edge weight denoting topological overlap between connected nodes, node size denoting within-module connectivity (F) Relationship with cohorts, showing mean level of module eigengenes.

Traditionally, MS-based proteomics is a low throughput technique in the range of 10-20 samples/day when nano-flow UPLC is employed to maximize sensitivity (Aebersold and Mann, 2003). In this study we developed a high throughput quantitative proteomics workflow, using a combination of Evosep One HPLC and Bruker timsTOF Pro on a total of 2622 plasma samples from 1612 individuals (Figure 1A) in one batch across 28 fully randomized acquisition plates to minimize assay variability between individuals and cohorts. In total we acquired 4553 LC-MS analyses including a pre-fractionated, super-depleted (Beer et al., 2017) master pool at 100 samples/day in data dependent acquisition mode (DDA-PASEF). The complete dataset comprises 250 million MS/MS spectra matching to a total of 2782 protein groups. In addition, we injected two non-depleted master pool samples every 24 cohort sample injections for subsequent identification transfer (“match between runs”, STAR methods) and quality control (Figure S1).

In addition to the data sparsity common to MS, the highly heterogeneous sample composition and long acquisition process introduced potential variation. We customized data pre-processing to minimize potential technical bias and maximize comparability between samples (workflow Figure S1A, STAR Methods). We analyzed raw protein intensities derived from FragPipe (STAR Methods), identifying 291 proteins reliably detected (in ≥50% samples) in at least one biological group, and removed 32 samples with few proteins detected and 22 proteins affected by cell residue contaminations in the plasma (Figure S1B-E; STAR Methods). We used Variance Stabilizing Normalization to normalize raw intensities and account for systematic bias, and applied K-Nearest-Neighbors to impute missing values based on the most similar proteins (for 170 proteins detected in ≥60% of the samples) or imputation by random draw from down-shifted normal distributions for the remainder. The processed data comprised 269 proteins in 2575 samples from 1598 patients (Figure 1A).

### An axis of severity across cohorts revealed by the proteome profile

We first sought to understand variation in plasma protein abundance and enrichment for biological processes across all cohorts. Reducing the dimensionality of the data, we found principal component (PC) 1 formed a sample gradient from HV and pre-operative, to post-operative and non-infectious critical illness, to sepsis and septic shock (Figure 1B). Across the patient and control cohorts, proteins with high positive loadings for PC1 included acute-phase (CRP, SAA1, SAA2, SERPINA1, SERPINA3, HP, C1RL), S100 pro-inflammatory (S100A8, S100A9, S100A12), innate immune or anti-bacterial (LCN2, LBP, USP15), and extracellular matrix (ECM) proteins (TNC, MMP2, COL1A2, COL6A1), while lipid transport protein APOM (Apolipoprotein M) had a high negative loading (Figure 1C). We identified protein clusters enriched for biological processes based on protein-protein interactions (Figure S1F).

We further analyzed functional groupings by deriving protein set enrichment scores using GSEA (STAR Methods). We analyzed the resulting matrix by principal component analysis and found that PC1 showed a gradient across sample cohorts with highest loadings for antimicrobial humoral response, cell chemotaxis, and positive regulation of response to external stimulus (towards sepsis/severe disease); and lipoprotein metabolic process (highest scores in non-sepsis controls). PC2 involved regulation of B cell activation, and Fc receptor signaling pathway (Figure 1D).

We also derived protein co-expression networks (STAR Methods), grouping 184 proteins into 16 co-expression modules that varied by comparator cohort and were enriched for acute-phase proteins (blue module, higher in sepsis), plasma lipoprotein assembly (yellow, higher in HV), platelet degranulation (brown, higher in sepsis) and immunoglobulins (black, purple and magenta, higher in sepsis and/or non-infected ICU patients) (Figures 1E, 1F, S1G).

### Sepsis is associated with a distinct plasma proteome profile

We proceeded to investigate differential protein abundance and pathway enrichment in sepsis. Here and subsequently, sepsis refers to the CAP/FP patients with sepsis admitted to ICU in the UK GAinS study unless stated otherwise. Across the six sepsis-comparator group contrasts in the discovery and validation cohorts (Figure 2A), we found 11 proteins differentially abundant in all contrasts, all with highest abundance in sepsis. These involved the acute phase response (CRP, LCN2, SERPINA1, HP), extra-cellular matrix (MMP2, COL6A1, TNC), protection in tissue damage (SERPINA1, HP, TNC), neutrophil function (LCN2, MMP2, SERPINA1, S100A9, S100A12), cytokine production (LCN2, MMP2, USP15, SERPINA1, HP, TNC, S100A12), and galactose metabolism (B4GALT1) (Figures 2B, 2C, S2A).

**Figure 2.**
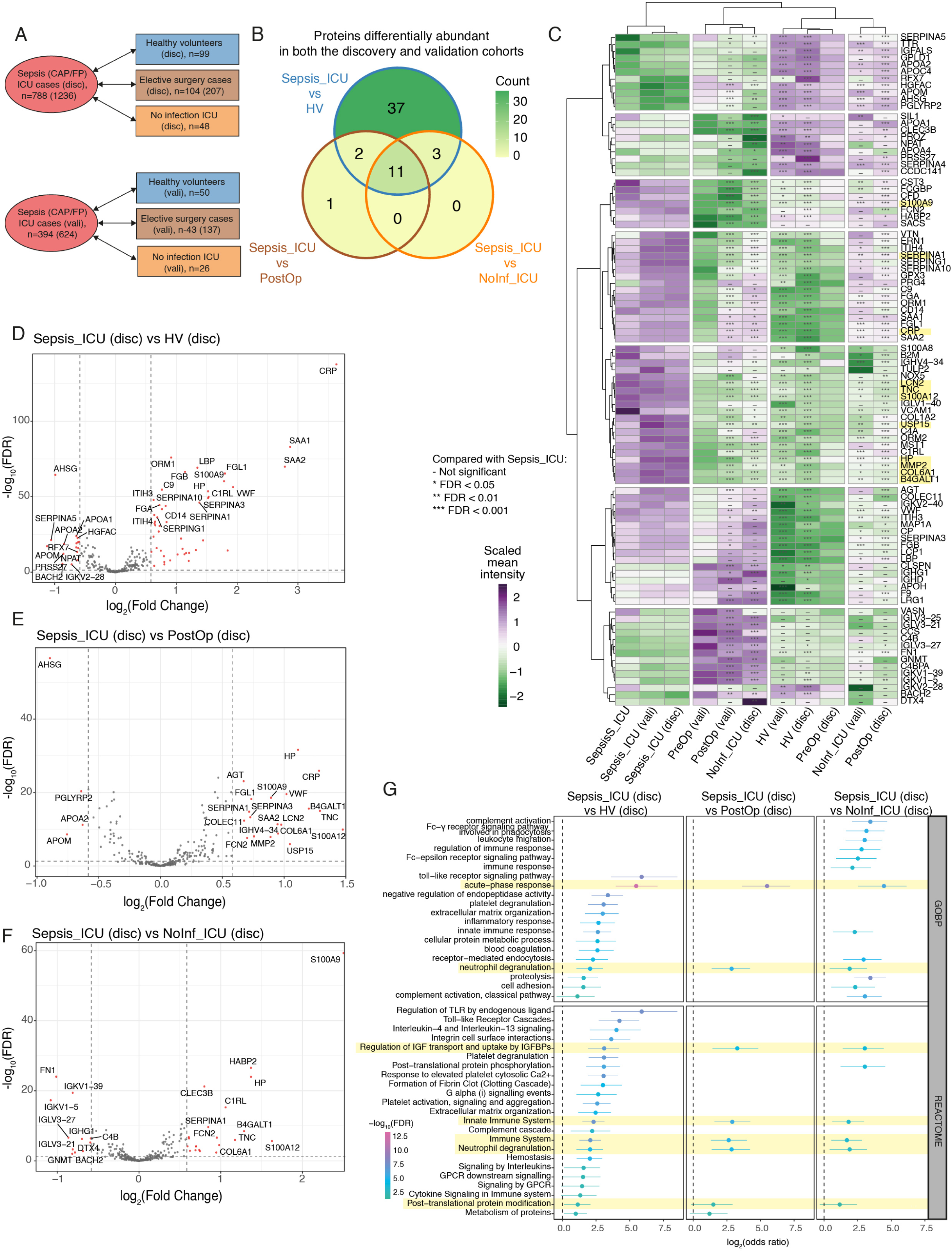
Sepsis-specific proteomic response. (A) Contrasts made. n, number of individuals (samples). (B) Venn diagram of differentially abundant (DA) proteins overlapping between contrasts. (C) Summary heatmap of mean protein abundance in sepsis and comparator groups, scaled by row. 94 proteins DA in any of the six contrasts in (A) are included, with significance level shown; 11 proteins DA in all contrasts shaded yellow. Only the first available samples per sepsis patient included. (D-F) Volcano plots of differential protein abundance for discovery cohort contrasts. Significantly different proteins (FDR<0.05 and |FC|>1.5) shown in red. (G) Pathway enrichment of DA proteins. Terms significantly enriched in all discovery cohort contrasts shaded in yellow. Horizontal bars indicate 95% confidence intervals of log_2_(odds ratio).

Compared with HV, we found 53 proteins were differentially abundant (FDR<0.05, |FC|>1.5) in both the discovery and validation cohorts (Figures 2B, 2D, S2B). Proteins more abundant in sepsis were implicated in the acute-phase response (CRP, SAA1, SAA2), coagulation process (VWF, FGB, FGA), and immune or immune-regulatory functions (LBP, S100A9, FGL1, ORM1, CD14), while reduced levels of apolipoproteins, alpha-2-HS-glycoprotein (AHSG), hepatocyte growth factor activator (HGFAC), plasma serine protease inhibitor (SERPINA5), transthyretin (TTR, inhibited by inflammation), and transcription regulator protein BACH2 (regulates apoptosis and adaptive immunity) were found.

We further compared sepsis with post-operative samples from elective surgery (X-MINS, BIONIC cohorts) reflective of a sterile inflammatory response and found 14 proteins including HP (haptoglobin), TNC (tenascin), B4GALT1, and S100A12 to be consistently differentially abundant. Of these, only one (ficolin-2, FCN2 which activates the lectin complement pathway (Matsushita, 2010)) was not also differentially abundant in the sepsis vs HV contrast (Figures 2B, 2E, S2C).

When we compared sepsis ICU patients to non-infected ICU patients, 14 proteins were differentially abundant in both the discovery and validation cohorts including S100A9, HP, SERPINA1, and B4GALT1 (Figures 2B, 2F, S2D) of which only C1RL (prohaptoglobin serine hydrolase), SAA2, and IGHV4-34 (BCR Ig) were not differentially abundant when sepsis was compared with post-operative samples.

We then sought to identify biological pathways consistently altered in sepsis. In the discovery cohort, sepsis differed from all comparator groups in acute-phase response, neutrophil degranulation, regulation of IGF (insulin-like growth factor) transport and uptake by IGF binding proteins, innate immune system, and post-translational protein modification (Figure 2G). Immune and metabolic processes that differed in sepsis vs HV and were not differential between sepsis and post-operative patients included TLR signaling, clotting, and IL-4 and IL-13 signaling. All these enriched terms were replicated in the validation cohort.

### Specific plasma protein subsets associate with sepsis severity, clinical covariates, source, and progression

We then investigated whether specific plasma proteins were associated with particular clinical features of the sepsis response, combining samples from the sepsis discovery and validation cohorts. We first analyzed overall variance in the proteome within sepsis patients. The largest component of variance, PC1, showed significant positive correlations with features relating to illness severity including blood lactate, Sequential Organ Failure Assessment (SOFA) scores, presence of shock, and mortality; and significant negative correlations with blood pressure, platelets and lymphocyte proportion (Figures S3A, 3B). In terms of individual plasma protein abundance, we identified a protein set (including PTGDS, B2M, CFD, LCN2, VWF, COL6A1, USP15, MMP2, COL1A2, CD14, PLTP and CRP) highly correlated with clinical variables reflecting more severe illness, including total SOFA, APACHE, occurrence of shock or renal failure, and prothrombin time, while a second set (including SERPIND1, C3, APOA1, HRG, KNG1, and VTN) had strong negative associations (Figure 3A).We found five proteins (CRP, LCN2, USP15, COL1A2, MMP2) significantly more abundant in patients with FP compared to CAP (Figures 3A, 3B). Within CAP, bacterial compared with viral infections showed higher abundance of LCN2 (Lipocalin-2) which limits bacterial growth by sequestering iron-containing siderophores (Shields-Cutler et al., 2016).

**Figure 3.**
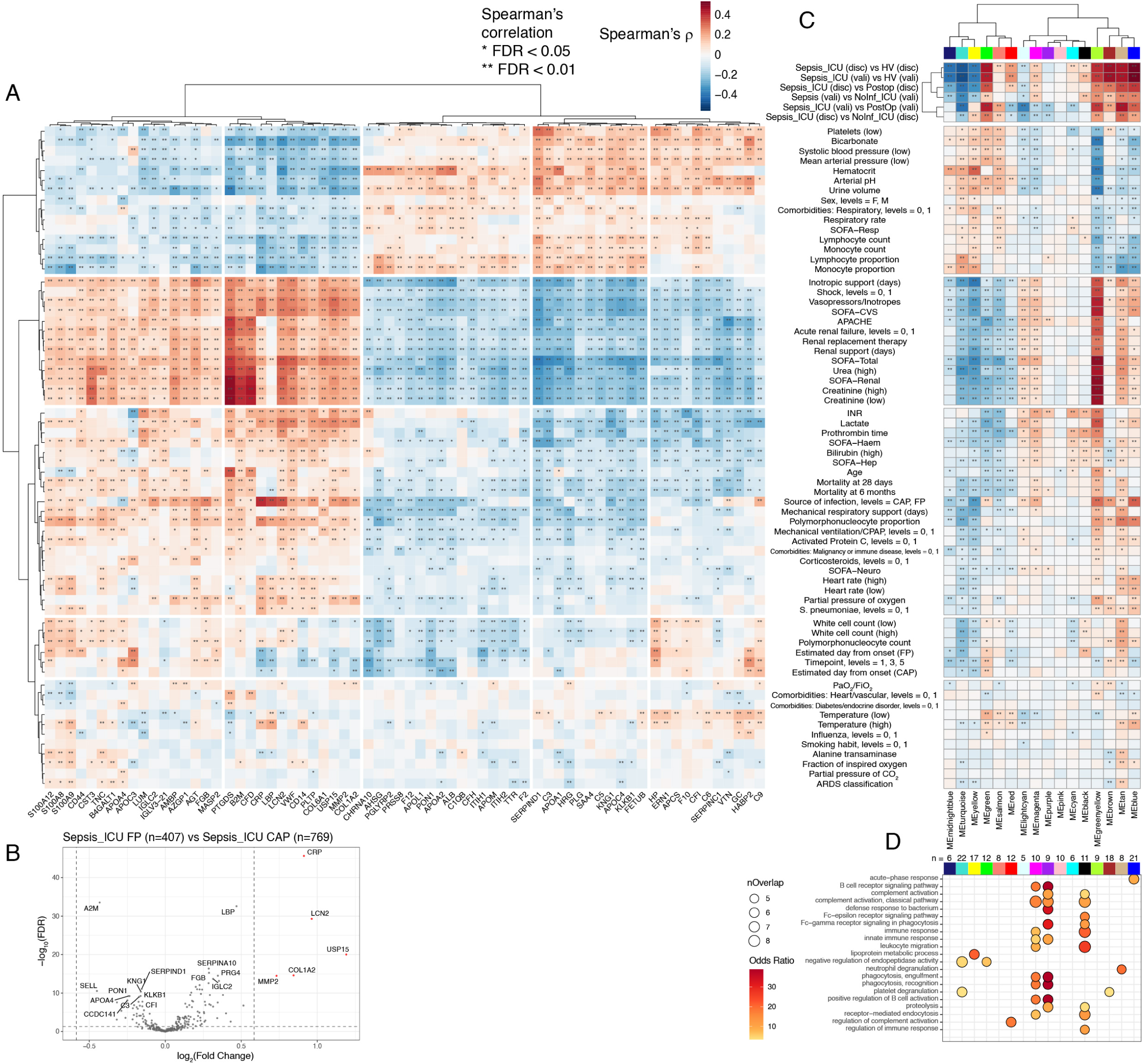
Variation within sepsis plasma proteome. (A) Heatmap of correlations between clinical characteristics (Table S1) and protein abundance (FDR<0.05, n=68 proteins shown), using first sample per patient (sepsis n=1182). Level 0/1 indicates absence/presence of trait. (B) Differential protein abundance for sepsis due to fecal peritonitis (FP) vs community acquired pneumonia (CAP) (using first available sample). (C) Heatmap of Spearman’s correlation between co-expression module eigengenes, group contrasts, and clinical variables. Row order aligned to (A). (D) Balloon plot of pathway enrichment for module member proteins (module size shown as n) using GOBP annotations.

Using the protein co-expression modules identified from all cohorts (Figures 1E, 1F; STAR Methods) we found specific modules were significantly correlated with comparator group contrasts and with specific clinical variables (Figures 3C, 3D, S3C). For example, the blue module, enriched for acute-phase response proteins, positively correlated with sepsis in all comparator contrasts but did not associate with mortality, showed modest association with level of organ dysfunction and the strongest association with high temperature. The tan module (contains S100 family proteins, enriched for neutrophil degranulation) showed a positive correlation in all comparator group contrasts with sepsis and a moderate positive correlation with severity features, high temperature and acute respiratory distress syndrome. The greenyellow module comprising many extracellular matrix proteins, while also positively correlated with sepsis, showed a stronger positive correlation with severity measures, together with renal impairment, lymphopenia, low temperature, increased mortality and increased age. The red module (complement activation) was associated with sepsis but correlated with less severe disease. Lipoprotein metabolic processes, reflected by the yellow module, were negatively correlated with sepsis, positively correlated with lymphocyte count, hematocrit, and a negative correlation with severity scores, renal impairment, mortality and mechanical ventilation.

Among sepsis patients with serial samples, 12 of 16 co-expression modules showed a change between ICU admission, Day3 and/or Day5 using paired samples (Figure S3D; Table S2). The blue (acute-phase), red (complement activation) and greenyellow (ECM) modules showed a consistent decrease over time from admission with the change in greenyellow module eigengene from Day1 to Day5 positively correlated with the change in SOFA total score (FDR=0.00052, rho=0.33 Spearman).

### Sepsis subphenotypes can be identified from the plasma proteome

We next sought evidence for sepsis subphenotypes from the plasma proteome. Consensus clustering on the protein intensities for the sepsis discovery cohort (Figure 1A) using all timepoints identified three subgroups, which we denote as Sepsis Plasma proteome-based Clusters (SPC1/2/3), and which represented the optimal cluster stability and number shown by cumulative distribution of the consensus index (Figures 4A-C, S4A; STAR Methods).

**Figure 4:**
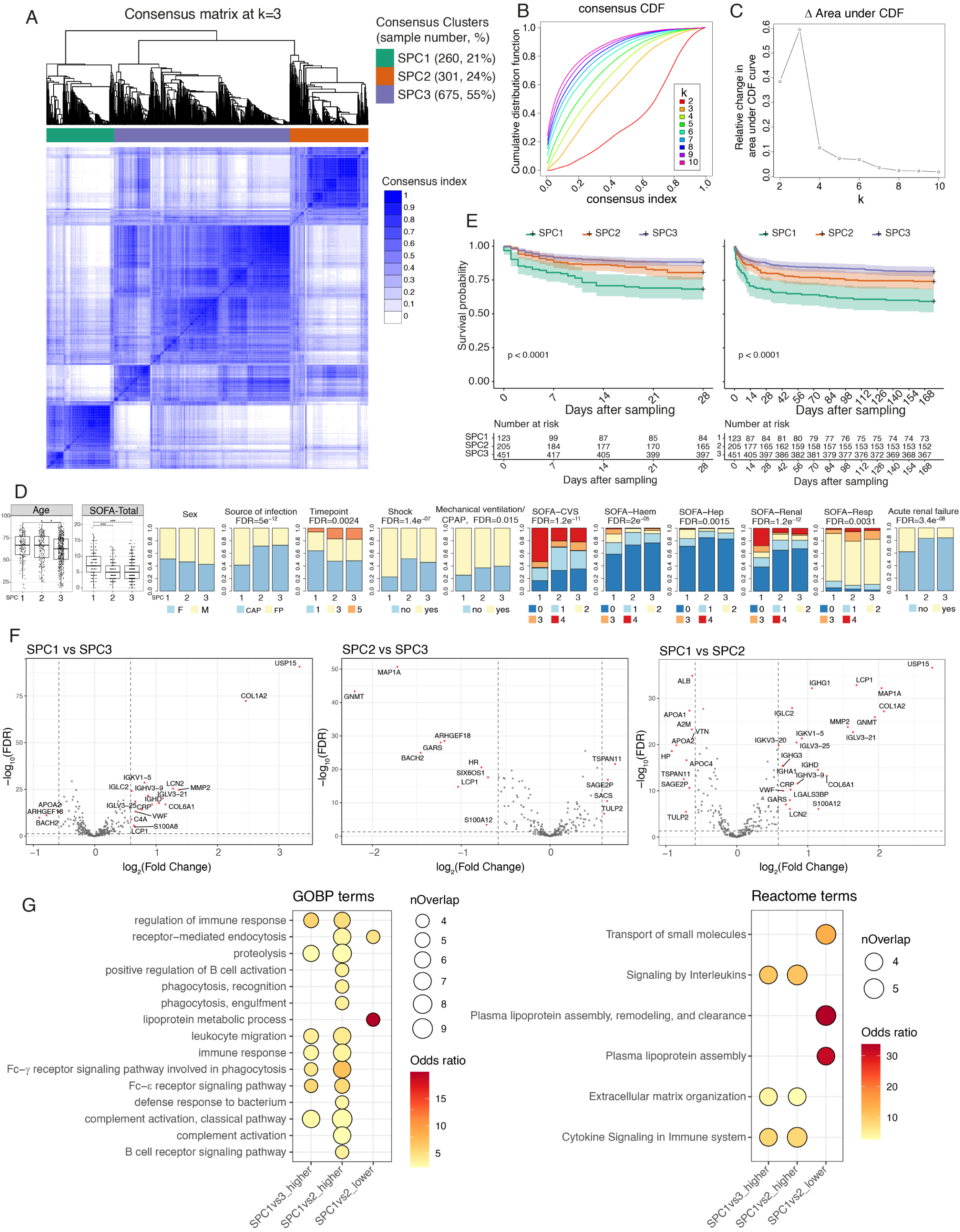
Sepsis Plasma proteome-based Clusters (SPC) in discovery cohort. (A-C) Consensus clustering (n=1236 samples, from 788 patients). (A) Cluster dendrogram and heatmap of consensus index (frequency of the sample pair being in the same cluster) for a cluster number of 3 with % of samples for each SPC noted (B) Cumulative distribution function (CDF) curves of the consensus index for increasing cluster number (k). (C) Relative change in area under CDF with k. (D) Bar plots of representative clinical variables between SPC (x-axis). Bar height represents proportion of each level. Age: SPC1 median 67 yrs (IQR 56-77), SPC2 67 (53-77), SPC3 63 (51-74) (Kruskal-Wallis FDR=0.014). Total SOFA score: median SPC1 7 (IQR 5-10), SPC2=SPC3 5(3-7), Dunn’s FDR SPC1vs2=2.3×10^-10^, SPC1vs3=1.2×10^-11^). Shock: SPC1=77%, SPC2=49%, SPC3=54% (χ^2^ FDR=1.4×10^-7^). Renal failure: SPC1=38%, SPC2=17%, SPC3=16% (χ^2^ FDR=3.4×10^-8^). Proportion FP (of FP+CAP patients): SPC1=59%, SPC2=28%, SPC3=27% (χ^2^ FDR=5.0×10^-12^). (E) Kaplan-Meier survival curves by SPC. For patients with multiple samples (Day1/3/5) cluster assignment from latest available sample used. Global p-values from log-rank tests; shading 95% confidence intervals. (F) Differential protein abundance between SPC. (G) GOBP and Reactome terms enriched in differentially abundant proteins. Only contrasts with significant terms detected shown.

We then investigated the clinical correlates of these clusters (Figures 4D, S4B; Table S3). Using the SPC assignments for the first available sample for each patient following ICU admission, we found that SPC1 comprised a more clinically severe subset of patients, with higher SOFA scores (except for respiratory SOFA) and more frequent occurrence of shock and renal failure (Figures 4D, S4B). SPC1 patients had significantly higher mortality than SPC2 and SPC3 at both 28 days (SPC1 vs 2+3 Hazard Ratio (HR) (95% CI) =2.5 (1.7-3.7), p=1.3×10^-6^) and 6 months (HR=2.3 (1.7-3.2), p=5.4×10^-7^) after sampling (Figure 4E). SPC3 patients were significantly younger than those in the other two clusters. SPC1 was enriched for FP patients and earlier time point samples. Restricting to CAP patients, we found those clustered as SPC1 had worse respiratory function and required more respiratory support, indicating that SPC1 identifies more severely ill patients after accounting for the original source of sepsis.

To confirm that the clustering was not biased by relatedness between serial samples from the same individual, we repeated our analysis of all discovery cohort patients using only the first available sample for each patient. Consensus clustering showed a robust three-cluster structure (88.7% of samples assigned as before). The first-sample cluster with the largest overlap with SPC1 also had significantly higher mortality than the other two clusters combined (HR (95% CI)=2.3 (1.6-3.2) at 28-day).

We then sought to understand the proteomic differences between SPCs. Compared to HV, more plasma proteins were differentially abundant in SPC1 (81, 74, and 55 proteins for SPC1, 2, and 3 sepsis patients respectively) including immunoglobulins and apolipoproteins specific to SPC1, enrichment for phagocytosis and positive regulation of B cell activation in SPC1, and lower levels of immunoglobulins in SPC2 (Figure S4C). Restricting the differential abundance analysis to between SPCs (Figures 4F, S4D), we found the proteins most abundant in SPC1 vs SPC3 were Ubiquitin carboxyl-terminal hydrolase 15 (USP15) and Collagen type I α-2 chain (COL1A2). Few proteins showed lower abundance in SPC1; these included apolipoprotein A-II (APOA2), transcription regulator protein BACH2, and Rho activator ARHGEF18. Compared to SPC2, SPC1 showed higher abundance of immune function proteins and immunoglobulins and lower albumin, apolipoproteins, haptoglobin, and cell adhesion mediators. SPC2 showed mostly lower protein abundance vs SPC3, specifically of intracellular proteins such as microtubule-associated protein 1A (MAP1A), metabolic regulator glycine N-methyltransferase (GNMT), and BACH2. Comparing SPC1 to SPC2 or to SPC3, we observed relatively greater activity of immune pathways, including interleukin signaling, Fc-γ or Fc-ɛ receptor signaling, leukocyte migration, complement activation, and ECM organization (Figure 4G).

### Proteomic patient subgroups are reproducible and involve specific pathways and biomarkers

In order to validate and further characterize these subgroups, we developed SPC prediction models. An elastic net model with 181 predictors performed best in terms of test-set accuracy (91.4%, Figure S5A). We applied this model to the sepsis validation cohort and replicated the associations with mortality (Figure 5A) and measures of severity including lactate, cell counts, vasopressor and renal support, and SOFA scores (Figures 5B, S5B). Differential abundance and pathway enrichment analysis between the validation cohort clusters and HV showed strong concordance with the discovery cohort (Figure S5C-E). We further tested cluster prediction based on a small number of informative protein biomarkers. We derived a new minimal elastic net model with 8 predictors (USP15, COL1A2, APOA2, MAP1A, GNMT, TSPAN11, LCP1, ALB), which successfully classified 79.5% of the test set, with a 72.7% sensitivity for SPC1.

**Figure 5:**
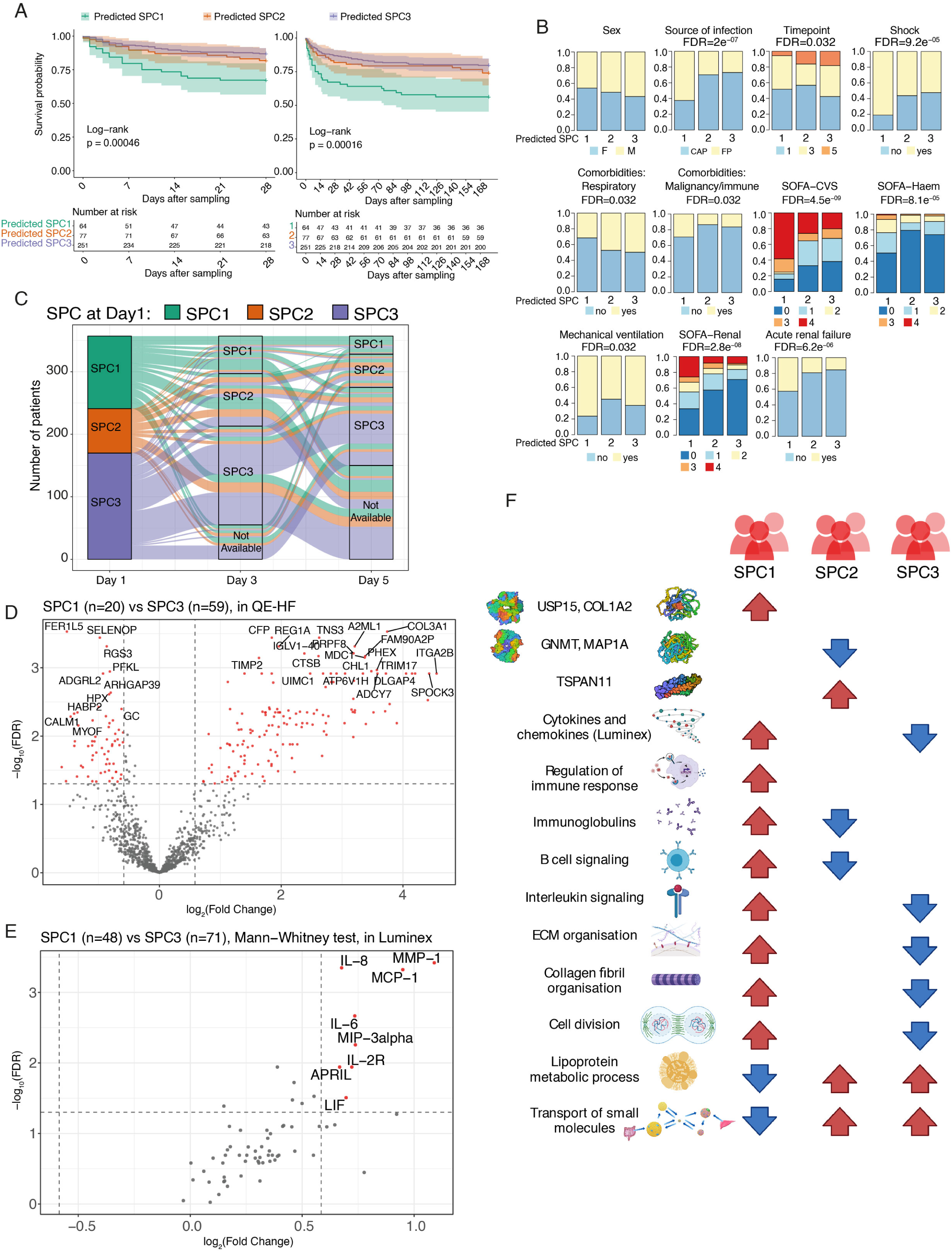
Validation and molecular characteristics of Sepsis Plasma proteome-based Clusters (SPC) (A, B) Sepsis validation cohort (A) Kaplan-Meier survival curves (n=392). (B) Bar plots comparing categorical clinical variables between SPC. (C) SPC movement Day1-5 of ICU admission (n=346 sepsis patients with a Day1 and at least one subsequent sample available, discovery and validation cohorts. Flow widths proportional to number of patients with the corresponding SPC transition. (D-E) Differential abundance analysis between SPCs using QE-HF mass spectrometer (D) or Luminex immunoassay (E) in subsets of sepsis samples. (F) Summary of molecular characteristics for each SPC (see also Table S7). Red and blue arrows indicate higher or lower abundance of the corresponding proteins respectively.

We also investigated the transitions between clusters. Cluster membership was significantly associated with time following ICU admission (χ^2^ p=0.0012). Considering only the 526 patients with multiple time points available, 57.4% of patients changed group over time, most frequently from SPC1 to SPC3 suggesting a general trajectory of recovery (Figure 5C, Table S4). SPC assignment could therefore be useful not only for defining sepsis response state and prognostication, but also for therapeutic intervention (based on evidence of patient movement between SPCs) and monitoring disease progression.

The EvosepOne-timsTOF platform enabled us to profile unprecedented numbers of patient samples, but with limitations in the range of proteins that could be detected. We therefore employed two additional methods to more fully characterize these clusters in a subset of patients. Firstly, we profiled 148 sepsis samples from 100 patients on a QE-HF mass spectrometer after depleting 12 highly abundant proteins. This permitted measurement of many more proteins (1123 detected in ≥70% samples, STAR Methods). Recognizing power was greatly reduced due to the restricted sample size, and potential limitations associated with the depletion process, we identified 144 proteins with higher and 63 proteins with lower abundance in SPC1 vs SPC3, and no signal in the two contrasts with SPC2 (Figure 5D). Pathway enrichment analysis again highlighted immune response pathways, ECM organization and lipoprotein metabolism differentiating SPC1 and SPC3, along with IL-4 and IL-13 signaling, collagen organization and the cell cycle. Secondly, we used the Luminex immunoassay to investigate 65 cytokines and other signaling molecules in 204 samples from 146 patients with SPC assignments. At the first available timepoint, the majority of analytes measured had higher sampled median concentrations in SPC1 vs SPC3, with significantly increased chemokines MCP-1, IL-8, and MIP-3α; cytokines involved in B cell proliferation (APRIL, IL-6), and immune inhibitory functions (IL-2R, LIF), and the interstitial collagenase MMP-1 (FDR <0.05 and FC>1.5) (Figures 5E, S5F). The cytokine profile of SPC1 indicated greater activity in chemotaxis and IL-6 regulated pathways.

Overall, SPC1 was characterized by higher abundance of immune response proteins, including specific cytokines and immunoglobulins, and more collagen and ECM components in the circulating plasma, implying a greater degree of tissue damage in these patients (Figure 5F; Table S5). Lipoprotein metabolism and transport were comparatively downregulated in SPC1. SPC2 by contrast had lower levels of immunoglobulins and B cell signaling pathway proteins, whereas in SPC3 interleukin signaling and cytokine concentrations were relatively reduced.

### Integration of plasma proteome and leukocyte transcriptome reveals components contributing to the sepsis response

We next sought to maximize the informativeness of the sepsis plasma proteomics (MS) by integrating with paired white blood cell transcriptomics (RNAseq) for 837 samples (649 patients), using matrix decomposition (Hore et al., 2016) to derive 284 latent components. These each comprised vectors of scores (loadings) that indicate the contribution of individual proteins and/or genes linked by that component, with 76 components having significant contributions from proteins. We then tested for correlation or association of the 284 components with clinical severity, patient subgroup, etiology, and time point (Figure 6; Table S6). To illustrate the approach, for sex we found a single highly significant component (Figure S6A), but for other phenotypes several components were correlated.

**Figure 6:**
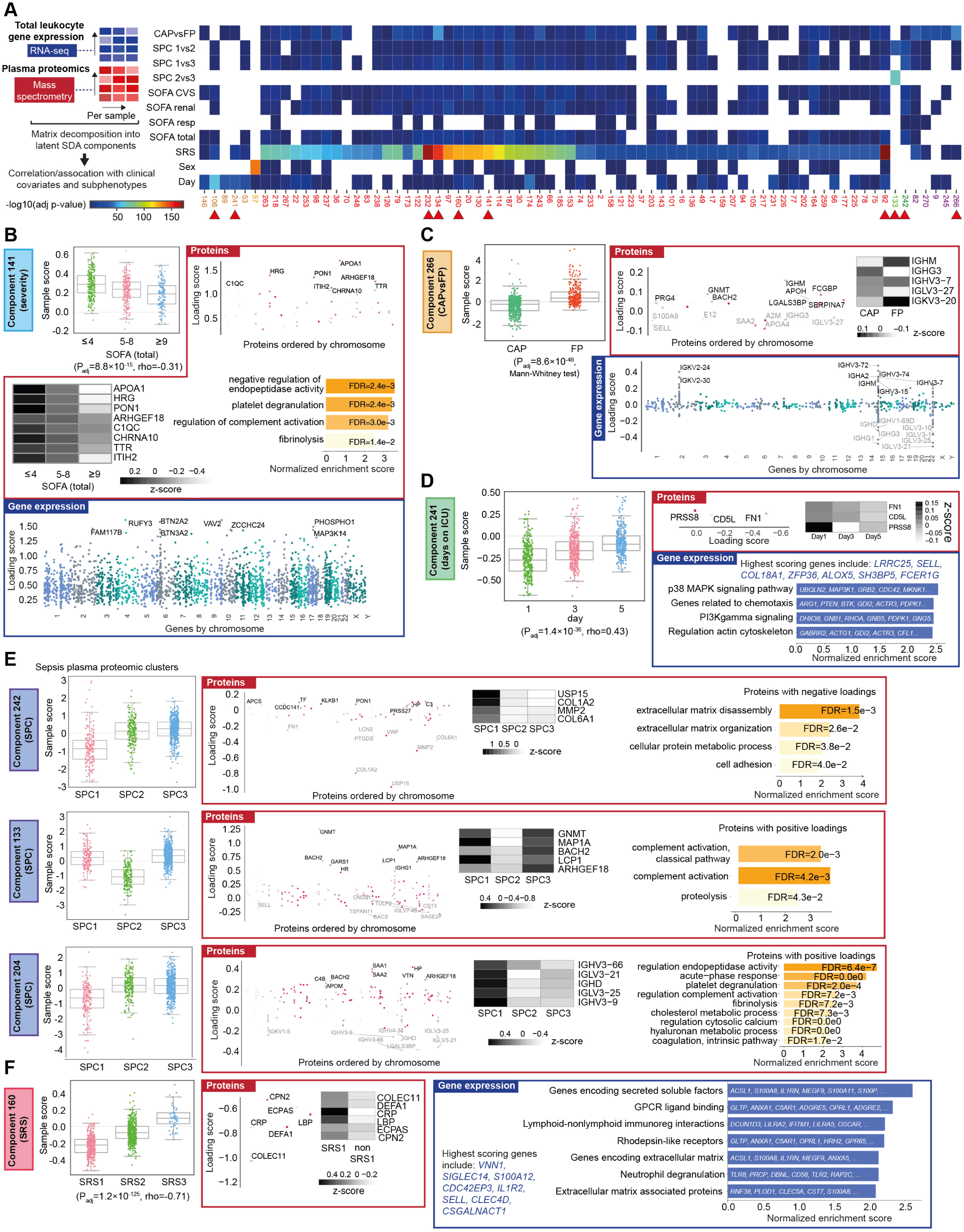
Integration of plasma proteomics and leukocyte transcriptomics. Matrix decomposition of proteomic and transcriptomic data (n=837 samples from 649 sepsis patients). (A) Heatmap showing 75 latent components with highest significance in correlation tests/group contrasts. Red arrowheads indicate components described in text. (B-F) For representative components: sample loading scores by group (boxplots), protein or gene loading scores for those significantly contributing to the component (posterior inclusion probability >0.5), z-scores of representative proteins, and pathway enrichment of significantly contributing proteins or genes.

The two components (187, 164) most significantly correlated with disease severity (total SOFA scores) involved only differential gene expression and implicated metabolic and immune processes (Figures S6B, S6C). By contrast component 141 linked lower SOFA scores to genes enriched for HLA class II and proteins implicated in lipid biology (APOA1, PON1) and histidine-rich glycoprotein (HRG) (a proposed sepsis biomarker important in maintaining neutrophils in a quiescent state (Kuroda et al., 2018; Takahashi et al., 2021)) with overall enrichment for negative regulation of endopeptidase activity, platelet degranulation, and regulation of complement activation (Figure 6B).

In terms of components significantly correlated with source of sepsis (CAP vs FP), component 134 had only gene expression contributions, including neutrophil activation marker *CD177*, bone marrow kinase BMX (role in endothelial permeability in sepsis (Li et al., 2020)) and matrix metalloprotease MMP9 (ECM degradation, implicated as a sepsis biomarker (Lorente et al., 2009)) (Figure S6D). The second most significant component 266 linked contributions from multiple differentially expressed genes and cognate proteins for immunoglobulin variable and constant chains, and enrichment in receptor-mediated endocytosis (FDR=0.016) in FP (Figure 6C).

We also identified components correlated with time from ICU admission (Table S6). Component 106 linked differential gene expression and later timepoint, involving ephrin receptor (endothelial cell migration) *EPHB4*, cholesterol metabolism (*CYP27A1*), inflammation and tissue remodeling (*CHI3L1*), transferrin neutrophil granules (*LTF*) and integrin (*ITGB4*) but not proteins, while component 174 revealed genes contributing to early time points including T cell activation (costimulatory gene *TNFSF4*), transcriptional regulation (*FAM172A*, *MAML3*), fatty acid biosynthesis (*OLAH*) and glycolysis (*TPK1*) (Figure S6E,F). Component 241 had the third highest significance, with proteins associated involving PRSS8 (prostasin serine protease), CD5L (lipid synthesis, macrophage apoptosis), and FN1 (Fibronectin, cell adhesion and motility); and regulation of chemotaxis and signaling pathways indicated by gene expression loadings (Figure 6D).

The most significant components for SPCs all involved protein abundances only (Figure 6E). Component 242 had the most significant association with SPC1vs3 (P*_adj_*=9.2×10^-34^, Mann-Whitney test) and the second highest in SPC1vs2 (P*_adj_*=1.1×10^-19^), with contributions from proteins enriched for ECM and metabolism (USP15, COL1A2, MMP2, VWF). Component 133 showed the most significant association with both SPC1vs2 (P*_adj_*=3.3×10^-36^) and SPC2vs3 (P*_adj_*=1.0×10^-67^). Multiple proteins had high loading scores for this component, including GNMT, MAP1A, BACH2, and ARHGEF18. Component 204 differentiated SPC1 from SPC2 (P*_adj_*=2.3×10^-18^) and SPC3 (*P_adj_*=1.1×10^-22^), highlighting immunoglobulin variable chains that mostly had higher abundance in SPC1.

We have previously reported sepsis response signatures (SRS) from leukocyte transcriptomic datasets associated with outcome and differential response to therapy (Antcliffe et al., 2019; Burnham et al., 2017; Davenport et al., 2016), including patients with evidence of granulopoietic dysfunction involving specific neutrophil subsets, relative immune compromise and high mortality (SRS1) or changes involving T cell and adaptive immunity (SRS2) measurable as a quantitative trait SRSq (Cano-Gamez et al., 2022; Kwok et al., 2022). We found component 92 strongly correlated with lower SRSq (more similar to SRS2) (P*_adj_*=1.6×10^-172^, rho=0.78 Spearman) which included increased apolipoprotein APOF and differential RNA abundance including repressive chromatin regulator *SAMD1* and T cell regulator ligase *RNF114* (Figure S6G). Most components most strongly associated with SRS did not have contributions from proteins, for example, the second most correlated component 232 linked lower SRSq with higher expressed genes enriched for TCR signaling, cytokine signaling, and adaptive immunity (Figure S6H). Component 160 linked higher SRSq (SRS1) with contributions from proteins including COLEC11 (collectin-11, role in innate immunity and apoptosis), CRP, DEFA1 (neutrophil defensin 1), LBP (LPS-binding protein), ECPAS (proteasome adapter and scaffold protein), CPN2 (carboxypeptidase N subunit 2), and from genes enriched for secreted soluble factors, GPCR ligand binding, neutrophil degranulation and immunoregulatory interactions (Figure 6F).

Overall, the matrix decomposition analysis identified features of the plasma proteome and leukocyte transcriptome associated with sepsis severity, disease etiology, progression, and subphenotype; in some cases dominated by proteomic or transcriptomic features, and in others linking features across these domains.

### Transcriptomic and proteomic profiling reveal complementary but distinct sepsis subphenotypes and response states

We further explored the relationship between plasma proteome- and leukocyte transcriptome-derived sepsis subphenotypes by analyzing 1016 patients (1361 samples) where both SPC and SRS assignments (derived from gene expression, STAR Methods, (Cano-Gamez et al., 2022)) were available. Considering the first available time points, we found 70% of SPC1 patients were also assigned to SRS1 in the discovery cohort, compared to 37% and 34% in SPC2 and SPC3 (71%, 48% and 31% in validation cohort) (χ^2^ p<0.0001; Figures 7A, 7B). The subsequent movement between the clusters over ICU admission showed greater likelihood of transition from SPC1 and SRS1 to another state (Figures S7A, S7B).

**Figure 7:**
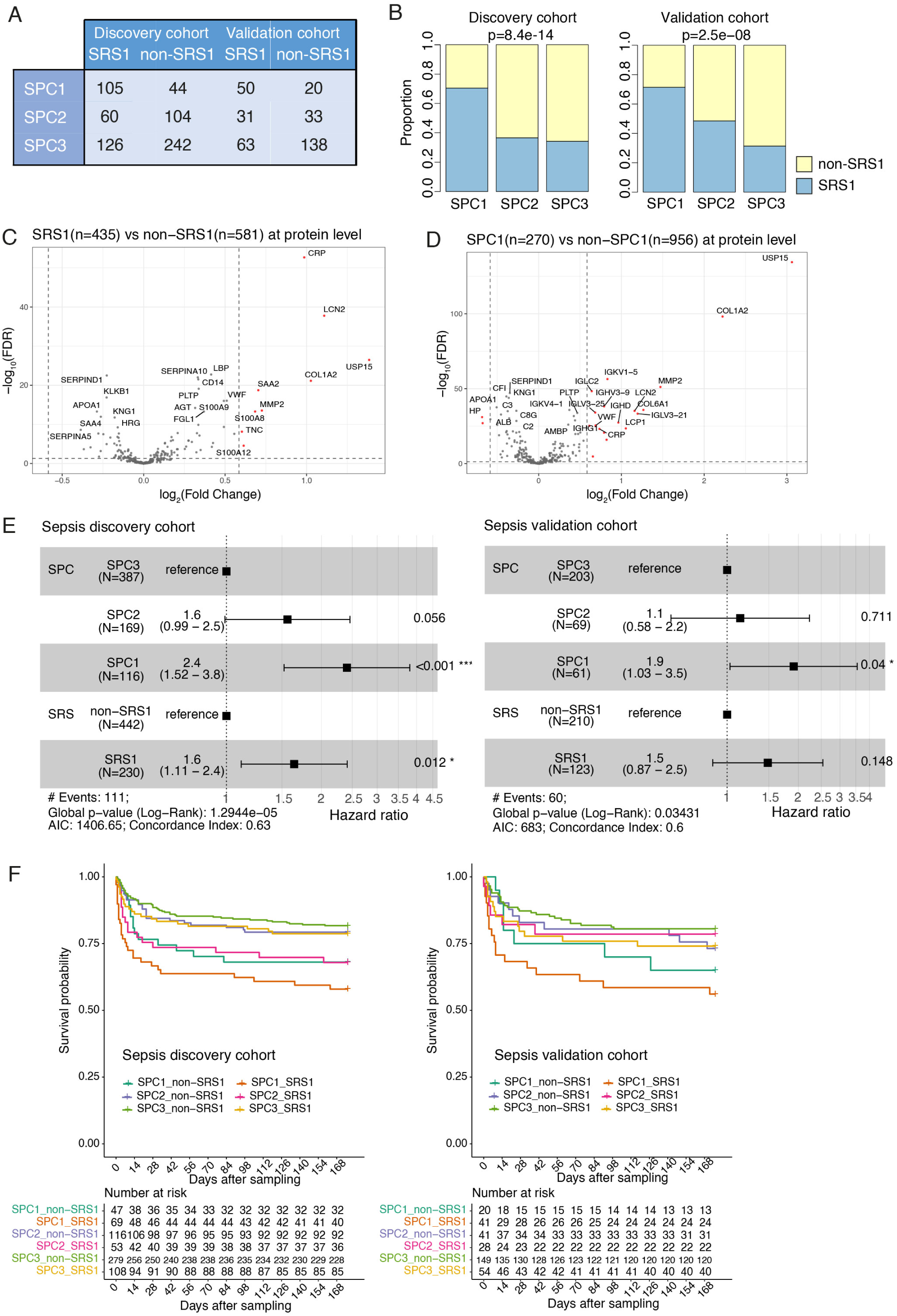
Interaction of proteomic (SPC) and transcriptomic (SRS) patient subgroups. (A-B) Overlap in SRS and SPC assignments in sepsis patients by (A) patient numbers and (B) proportions. First available samples per patient used. (C-D) Protein differential abundance analysis for SRS (C) and SPC (D). (E) Multivariate Cox proportional hazard regression on 28-day mortality considering both SPC and SRS assignments. Cluster assignments of last sample (within ICU Day1/3/5) per patient with both SRS and SPC assignments used. (F) Kaplan-Meier curves comparing survival at 6-months post-sampling by combined SPC and SRS assignments.

We identified differentially abundant proteins between SRS groups (Figure 7C), some of which overlapped with the proteins discriminating SPC1 from SPC2+3 (Figure 7D) including higher abundance of CRP, LCN2, USP15, COL1A2, SAA2, MMP2, S100A8, TNC and S100A12 in both SRS1 and SPC1. On the other hand, a set of immunoglobulins, HP and APOA2 differed only between SPC, and SAA2 only in the SRS contrast. Gene expression differences between SRS groups and SPC groups were strongly correlated (Figure S7C). Pathways enriched in the differential proteins and genes showed shared and specific features (Table S7), including cytokine signaling and innate immunity inferred from higher-abundance proteins in both SRS1 and SPC1, together with neutrophil degranulation and oxidation-reduction (upregulated) and adaptive immune response and T cell co-stimulation (downregulated) in both SRS1 and SPC1 from gene expression analysis. However, each contrast also had unique features, with MHC class II genes downregulated uniquely in SRS1, and IFN signaling and cell division terms only enriched in the SPC analysis (Table S7).

Finally, given SRS1 and SPC1 both associated with poor outcome (Figure 7E), we tested whether the two classifications can be combined to further inform risk stratification. We found the patients assigned to both SRS1 and SPC1 (∼11% patients) had the highest mortality rate of 33.3% at 28 days (31.7% in validation cohort), with Hazard Ratio (HR)=3.9 (95% CI 2.3-6.7) p<0.0001 (discovery) (HR=3.0 (1.5-6.0) p=0.002 (validation)) vs ‘SPC3 non-SRS1’ patients (∼43%) who had the lowest mortality of 10.4% (12.8% in validation cohort) (Figures 7F, S7D, S7E).

## Discussion

In this study we have mapped the human sepsis plasma proteome, applying an innovative mass-spectrometry based approach at scale to understand how sepsis differs from health, sterile inflammatory states and non-infected critical illness as well as individual variation in the sepsis response. The sepsis plasma proteome reflects mechanisms underlying the dysregulated host response to infection, as well as the wider consequences of organ dysfunction (reduced metabolism and excretion for example) and tissue injury. This offers the opportunity to identify new aspects of pathogenesis together with measures of organ dysfunction and disease severity.

We report evidence of specific proteins, co-expression modules and networks differentially abundant in sepsis involving innate immunity, acute-phase response, neutrophil function, cytokine production, lipometabolism, tissue damage protection and extra-cellular matrix organization. More severe illness was associated with specific proteins including PTGDS, B2M, CFD, LCN2, VWF, COL6A1, USP15, MMP2, COL1A2, CD14, PLTP and CRP, and modules enriched for S100 family proteins and extracellular matrix proteins (positive correlation), complement and lipoprotein metabolic proteins (negative correlation). We found dynamic changes over ICU admission, particularly involving acute-phase, complement and ECM related modules.

A key challenge relates to disease heterogeneity within sepsis, and the extent to which biological processes causing disease are shared or specific within critical illness more generally (Maslove et al., 2022). We and others have found evidence of sepsis subphenotypes based, for example, on clinical and lab variables (Seymour et al., 2019; Zador et al., 2019), inflammatory mediators (Fjell et al., 2013), and the leukocyte transcriptome (Baghela et al., 2022; Burnham et al., 2017; Davenport et al., 2016; Scicluna et al., 2017; Sweeney et al., 2018; Wong et al., 2019). Here we identify and validate three patient clusters from the sepsis plasma proteome, informative for response state, disease severity and outcome. SPC1 patients have the most severe disease and show differences in proteins related to phagocytosis, lipoprotein metabolism, interleukin signaling, chemotaxis, B cell activation and circulating immunoglobulins. These complement and add granularity to the leukocyte transcriptomics SRS groups. When considered together, patients in both SPC1 and SRS1 have the worst outcome, with shared features relating to cytokine signaling and innate immunity, neutrophil degranulation and oxidative metabolism.

Further work is needed to establish the mechanisms driving SPC and whether they are reflective of treatable traits. Animal and human studies, for example, already highlight lipoproteins as potential therapeutic targets in sepsis (Tanaka et al., 2020) and COVID-19 (Chidambaram et al., 2022). Additional work is also required to determine the maximally informative biomarkers appropriate for point of care testing, and whether these can be applied in combination with other ‘omic platforms and currently available clinical or laboratory variables to better stratify patients.

The increasing availability of multi-modal high dimensional data for clinical and molecular disease phenotyping requires innovative approaches to analyze and integrate such datasets (Graw et al., 2021; Rajczewski et al., 2022). Here, we have leveraged analytical strategies developed for transcriptomics to investigate protein co-expression networks and modules, and signatures of response. We have further shown how matrix decomposition allows integration of paired plasma proteomic and leukocyte transcriptomic data, demonstrating instances where these are linked in a specific component and informative for illness severity or disease subgroup. Reassuringly, we find a high concordance with proteins and processes identified from analysis of individual proteins, modules and latent components.

In this study we show that medium to high throughput proteomics across multiple large cohorts in a single batch is feasible on a single LC-MS platform. A simple semi-automatic sample preparation strategy in combination with the MS-based analysis of >2500 clinical, non-depleted plasma samples and further ∼2000 quality control and library samples at 100 samples/day now reaches throughputs employed by other proteomics technologies such as Olink or SOMAscan, but at significantly lower costs and sample usage. Although measured proteome depth is limited due to the extreme dynamic range of plasma protein abundance, we achieve good coverage of the acute phase proteome and markers of disease routinely quantified in single measurement assays such as ELISA. The throughput, cost effectiveness and robustness of modern LC-MS platforms mean this technology is now competitive with standard clinical practice measurements such as ELISA for absolute protein quantitation if heavy isotope labeled peptide standards are spiked into the clinical samples, marking a transition from a pure discovery tool towards a more clinical point of care application in the coming years. At the same time, LC-MS based proteomic analysis of plasma samples allows unprejudiced discovery analysis of novel biomarkers without compromising clinical, patient specific measurements. Future work will focus on streamlining sample and data processing workflows towards clinical certification and point of care use.

### Limitations of the study

Further work is needed to fully establish the complete sepsis proteome across a wide dynamic range of protein abundance and size, and differentiate protein variation (proteoforms) (Melani et al., 2022). Recent advances in Data Independent Acquisition (DIA) for MS would be compatible with high throughput proteomics platforms and offer future opportunities to increase depth and data completeness. It is not known to what extent individual organs and tissue beds contribute to the observed plasma proteome in sepsis and it would be valuable to quantify disease relevant tissue-specific proteomes, recognizing that obtaining tissue samples from the critically ill is very challenging. Application of co-expression analysis is limited by the relatively sparse nature of the proteomic data. Network-based methods such as similarity network fusion (Wang et al., 2014) and multi-layer patient similarity networks (Kivelä et al., 2014) may further facilitate inference of patient connectivity and subgroup detection.

## Data Availability

The raw and processed timsTOF mass spectrometry data are publicly available on the Proteomics Identification Database (PRIDE) (deposition in progress, accession ID TBC).
RNAseq gene expression data for GAinS study samples will be made available in the European Genome-Phenome Archive before publication. Microarray gene expression data for GAinS study samples are publicly available in ArrayExpress (E-MTAB-4421, E-MTAB-4451, E-MTAB-5273, and E-MTAB-5274).

https://www.ebi.ac.uk/arrayexpress/

https://ega-archive.org/

## Acknowledgements

Research was supported by the National Institute for Health and Care Research (NIHR) through the Comprehensive Clinical Research Network for patient recruitment. This research and participating researchers were funded by the Wellcome Trust (Wellcome Trust Investigator Award (204969/Z/16/Z) (J.C.K), core funding to the Wellcome Centre for Human Genetics (090532/Z/09/Z, 203141/Z/16/Z) and Wellcome Sanger Institute (206194, 108413/A/15/D), Medical Research Council (MR/V002503/1) (J.C.K., E.E.D., A.C.G.), CAMS IFMS 2018-I2M-2-002 (J.C.K.), China Scholarship Council-University of Oxford Scholarship (Y.M.), BJA/RCA Career Development Award (G.L.A.), British Oxygen Company (G.L.A.), British Heart Foundation (RG/14/4/30736 and RG/19/5/34463) (G.L.A.), NIHR Advanced Fellowship (NIHR 300097) (G.L.A.), NIHR Oxford Biomedical Research Centre (J.C.K., Oxford BioBank and Oxford Bioresource), NIHR Imperial Biomedical Research Centre (D.A., R.D., D.J.P.O, P.P., A.C.G., BIONIC), NIHR Research Professor award (RP-2015-06-018) (A.C.G), NIHR Research for Patient Benefit awards (MONGRAMS, PB-PG-0613-31073 and VANISH, PB-PG-0610-22350), Intensive Care Foundation Young Investigators Awards (TACE and MONOGRAMS), the National Institute of Academic Anaesthesia (BIONIC, BJA/RCoA Project Grant WKR0-2020-0019), European Society of Anaesthesiologists (BIONIC), and NIHR Academic Clinical Fellowship (ACF-2019-21-011) (H.D.T.). We thank participating patients, volunteers from Oxford Biobank (www.oxfordbiobank.org.uk), and laboratory members (of J.C.K., R.F. groups and the MS laboratory at the Target Discovery Institute Oxford led by Benedikt M. Kessler) for useful discussions. The views expressed are those of the author(s) and not necessarily those of the NHS, the NIHR or the Department of Health. For the purpose of Open Access, the author has applied a CC BY public copyright license to any Author Accepted Manuscript version arising from this submission.

## Author contributions

Conceptualization: J.C.K., R.F.

Data curation: Y.M., P.D.C.

Formal Analysis: Y.M., K.L.B., P.D.C., J.W., R.F.

Funding acquisition: R.F., J.C.K.

Investigation: Y.M., K.L.B., R.H., I.V., G.B.

Methodology: Y.M., P.D.C.

Project administration: J.C.K., R.F.

Resources: J.W., H.D.T., D.B.A., S.M.M., M.J.N., P.H., C.G., J.R., E.E.D., S.McK., R.D., D.J.P.O., P.P., F.K., A.C.G., G.L.A., C.J.H., J.C.K.

Software: A.N., F.Y.

Supervision: J.C.K., R.F.; supervision contributing cohorts: C.J.H., J.C.K., A.C.G., G.L.A., F.K., P.P.

Visualization: Y.M., K.L.B., J.C.K. I.V., R.F.

Writing – original draft: Y.M., K.L.B., J.C.K., R.F., I.V.

Writing – review & editing: all authors

## Declarations of interests

ACG has received consulting fees from AstraZeneca, unrelated to this project. Other authors declare no competing interests.

## Supplementary figure legends

**Figure S1.**
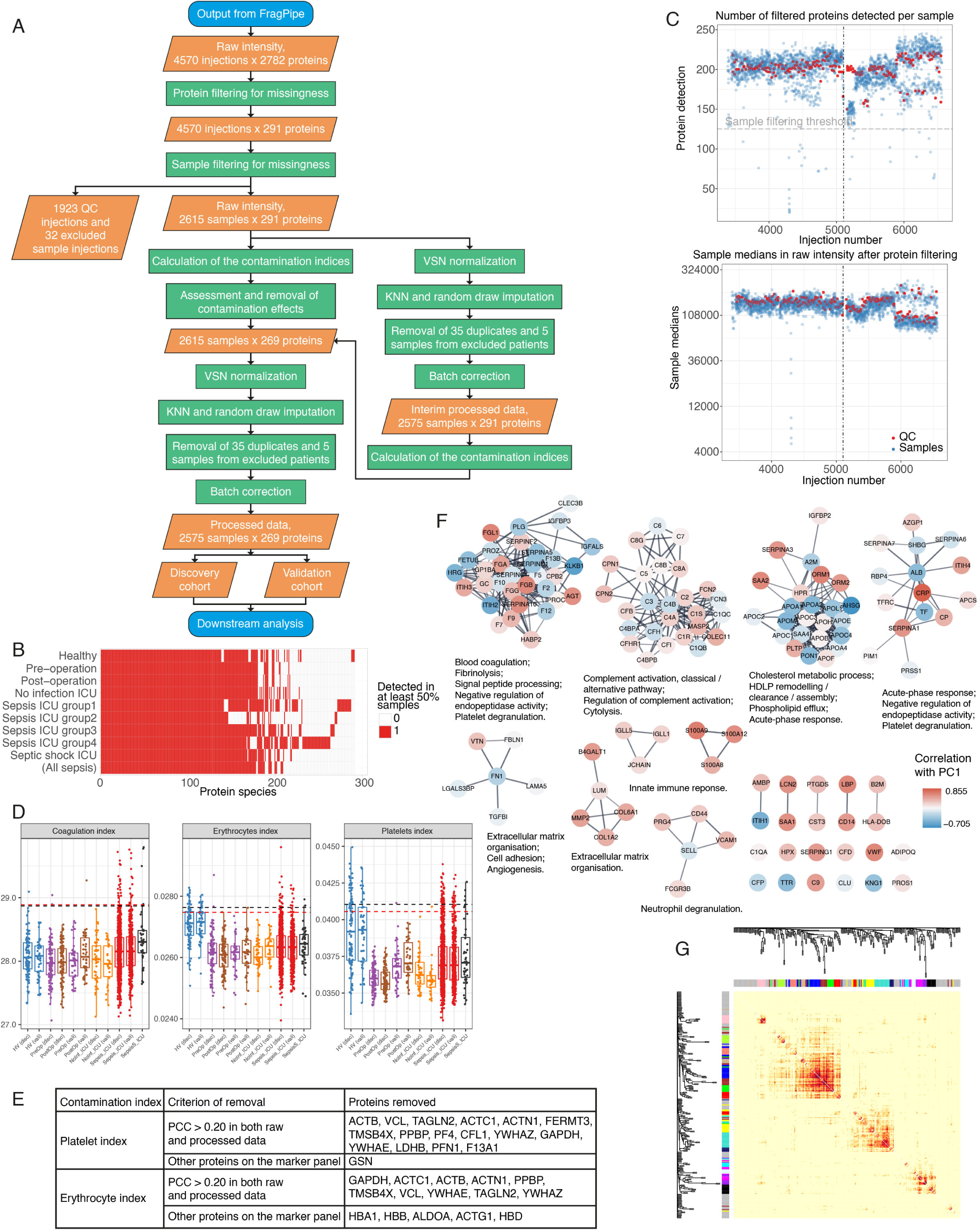
Mass spectrometry data pre-processing and protein co-expression modules, related to Figure 1. (A) Flowchart of the data pre-processing steps. Quality control (QC) injections refer to all the injections that did not come from individual samples. Samples from five patients were excluded for withdrawal of consent or having no clinical information available. (B) Heatmap showing whether the 291 remaining proteins post-filtering had ⩾50% detection in each of the groups, defined as described in STAR methods. The sepsis group as a whole is shown but was not used in protein filtering. (C) Scatter plots of numbers of filtered proteins detected and the median intensities in each injection along the acquisition process. “QC (system)” are the pools injected among the samples. Injection numbers correspond to the machine record in a chronological order, starting at 3390. Only the injection period containing the sample injections are plotted. The chromatographic column was blocked and changed at injection 5105 (vertical dash-dotted line). The horizontal grey dashed line indicates the sample filtering threshold (detection⩾125) used. (D) The contamination indices across the cohorts, calculated in the interim processed data. Horizontal red dashed lines indicate the sample outlier cut-off (mean + 2 s.d.) calculated in all samples, excluding HV in the erythrocytes index and platelets index; horizontal black dashed lines were calculated in the plotted samples. The two cut-offs are used only for visualization purposes but not for filtering samples. (E) List of proteins removed for being potential cell residue contaminations. (F) Clusters identified from protein-protein interaction (PPI) network of 141 measured proteins (STAR Methods) with node color mapped to Pearson’s correlation coefficients between the protein level and PC1 score across the samples, which correlated with disease severity. Within this network clusters enriched for coagulation and fibrinolysis, complement activation, cholesterol metabolic process, acute-phase response, ECM organization (two clusters), and neutrophil degranulation are shown. Gene ontology biological process (GOBP) terms enriched in the member nodes listed for each cluster. (G) The topological overlap matrix (TOM) based on protein co-expression in WGCNA. Rows and columns are the 269 protein species. Darker color in the heatmap represents a higher similarity between the two nodes. Gray in the color bar indicates the 85 proteins that were not co-expressed with other proteins in the dataset and should not be interpreted as a co-expression module.

**Figure S2.**
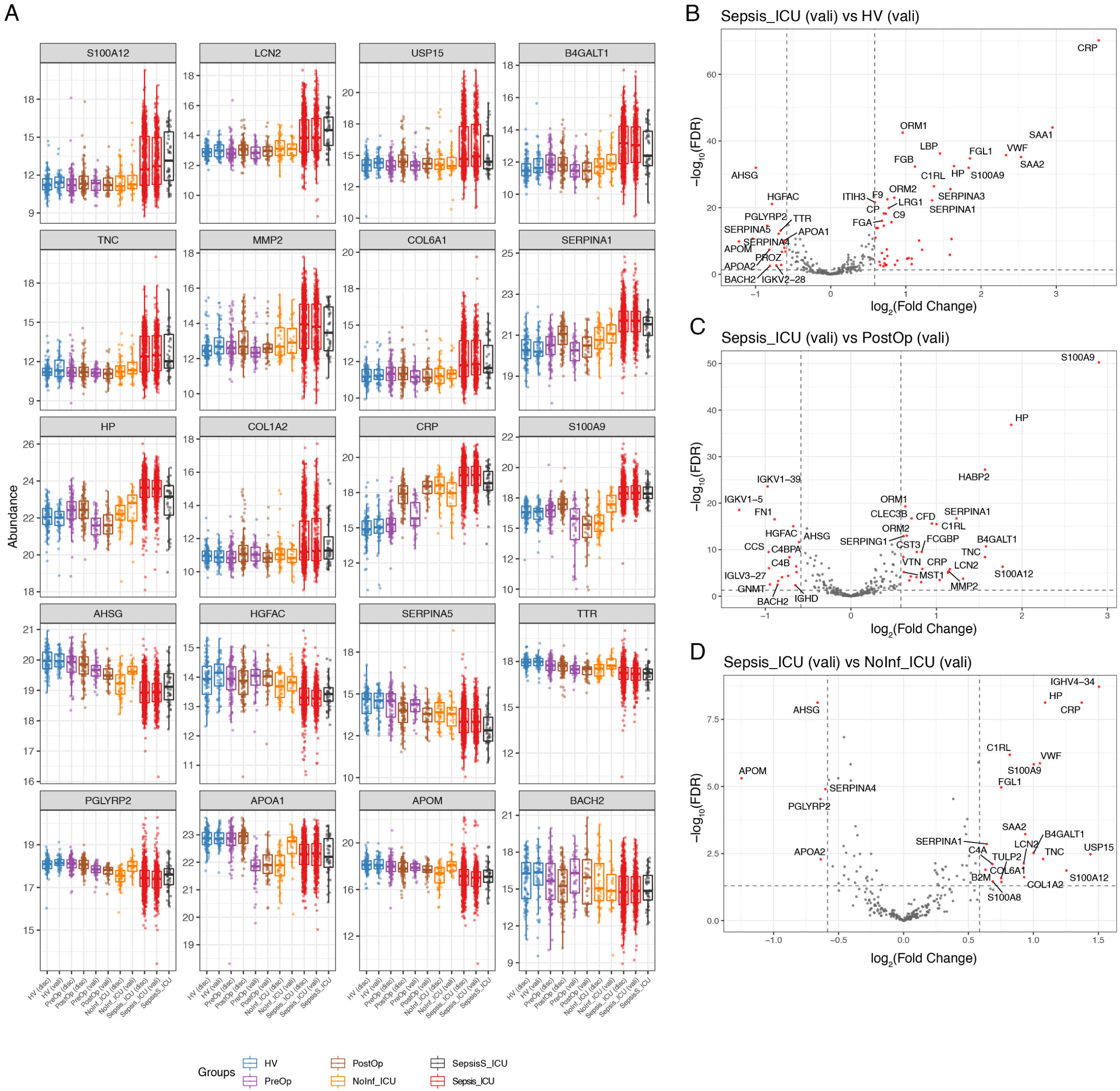
Sepsis-specific proteomic response, related to Figure 2. (A) Boxplots showing distribution of 20 representative proteins across the sepsis and non-septic comparator groups in the discovery and validation cohorts. (B-D) Volcano plots for the differential protein abundance for the sepsis-control contrasts (Figure 2A) in the validation cohort. Significantly different proteins (FDR<0.05 and |FC|>1.5) shown in red.

**Figure S3.**
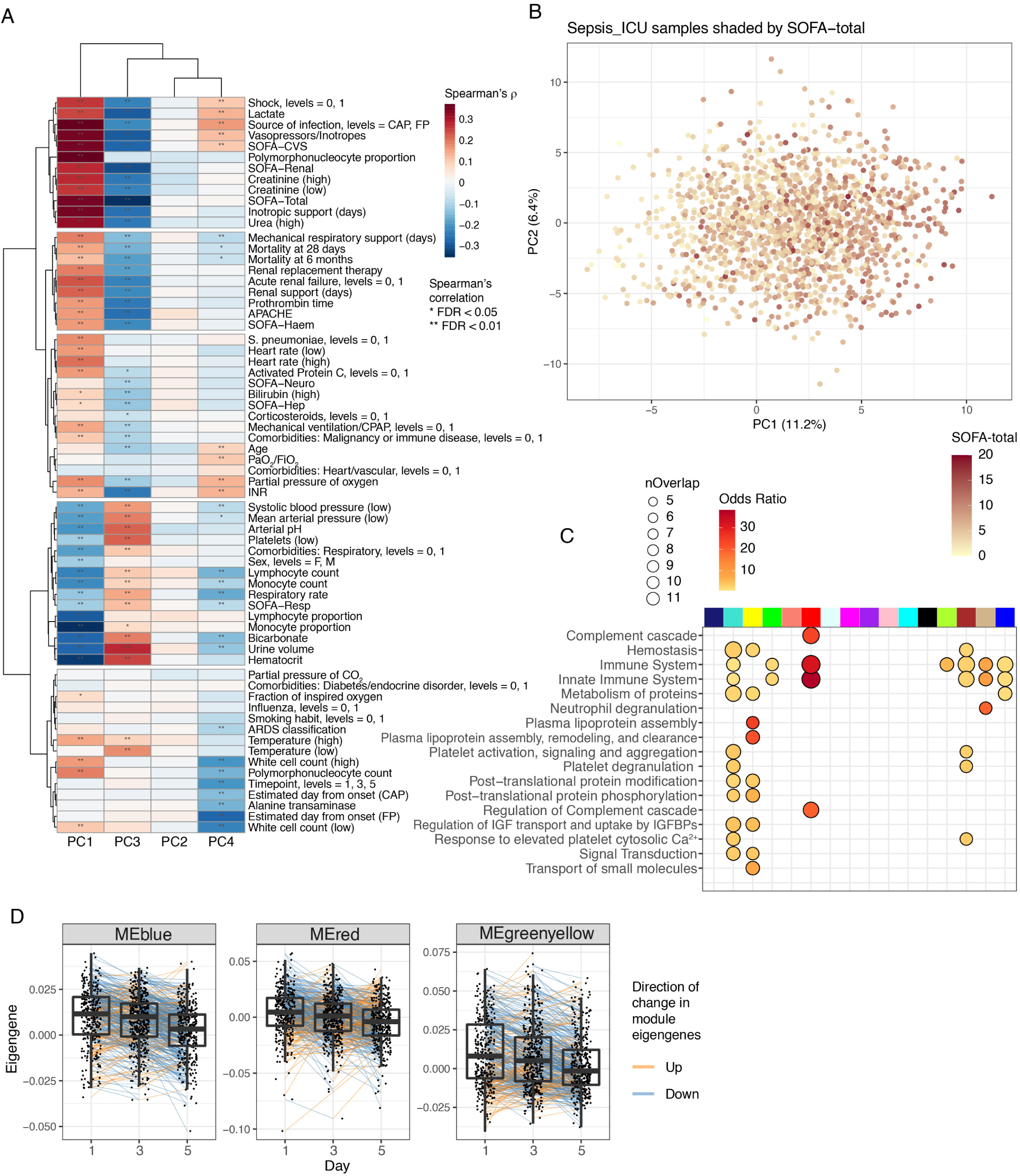
Variation within sepsis, related to Figure 3. (A) Heatmap of correlations between clinical characteristics in sepsis and the PC1-4 scores calculated using all samples. Correlations calculated in the first available samples after admission of Sepsis (CAP/FP) ICU patients. Level 0/1 indicates absence/presence. (B) Distribution of Sepsis (CAP/FP) ICU samples on PC1 and PC2 scores calculated using all samples, shaded by the SOFA-total scores on the day sampled. (C) Reactome pathway enrichment for member proteins of each co-expression module. Order of modules are as in Figure 3 B,C. (D) Module eigengene values on Day1/3/5 of ICU admission for Sepsis ICU samples. 1204 samples for 526 patients with at least two timepoints sampled are shown. Significance from group contrasts reduced to using paired samples are shown in Table S4.

**Figure S4.**
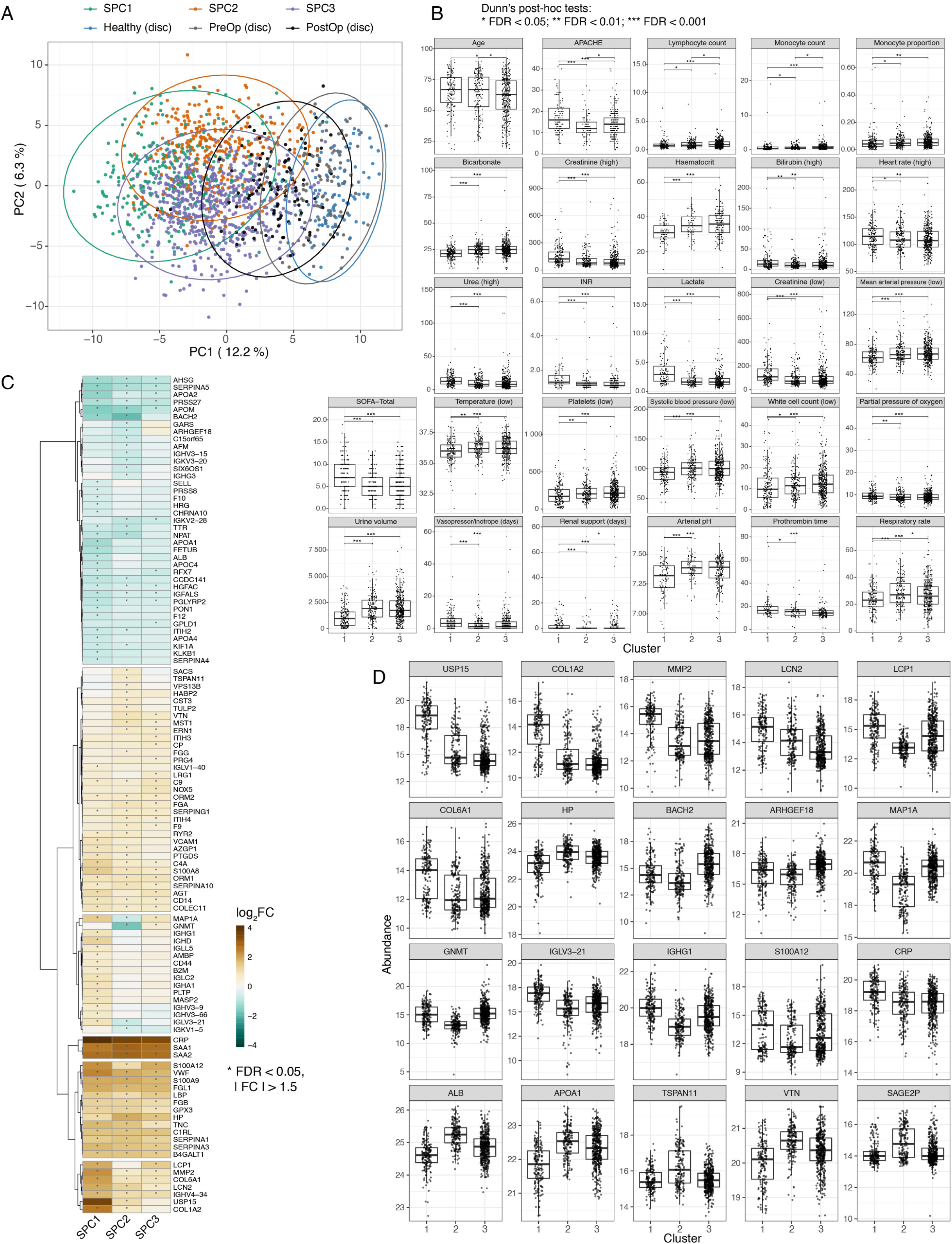
Characterization of the proteome-based patient subgroups in the discovery cohort, related to Figure 4. (A) Distribution of the sepsis subgroups on the first two principal components, plotted along with non-sepsis controls. Data ellipses are plotted for each group at a 95% confidence level assuming a multivariate normal distribution. (B) Box plots of numerical clinical variables that were significantly different between the subgroups (SPC). INR: international normalized ratio for prothrombin time. PaO_2_/FiO_2_: partial pressure of oxygen divided by fraction of inspired oxygen. (C) Heatmap of log fold changes comparing protein profiles of each cluster against HV, plotted for 111 proteins differentially abundant (* FDR<0.05 and |FC|>1.5) in any of the three contrasts. (D) Boxplots of representative proteins across the three clusters.

**Figure S5.**
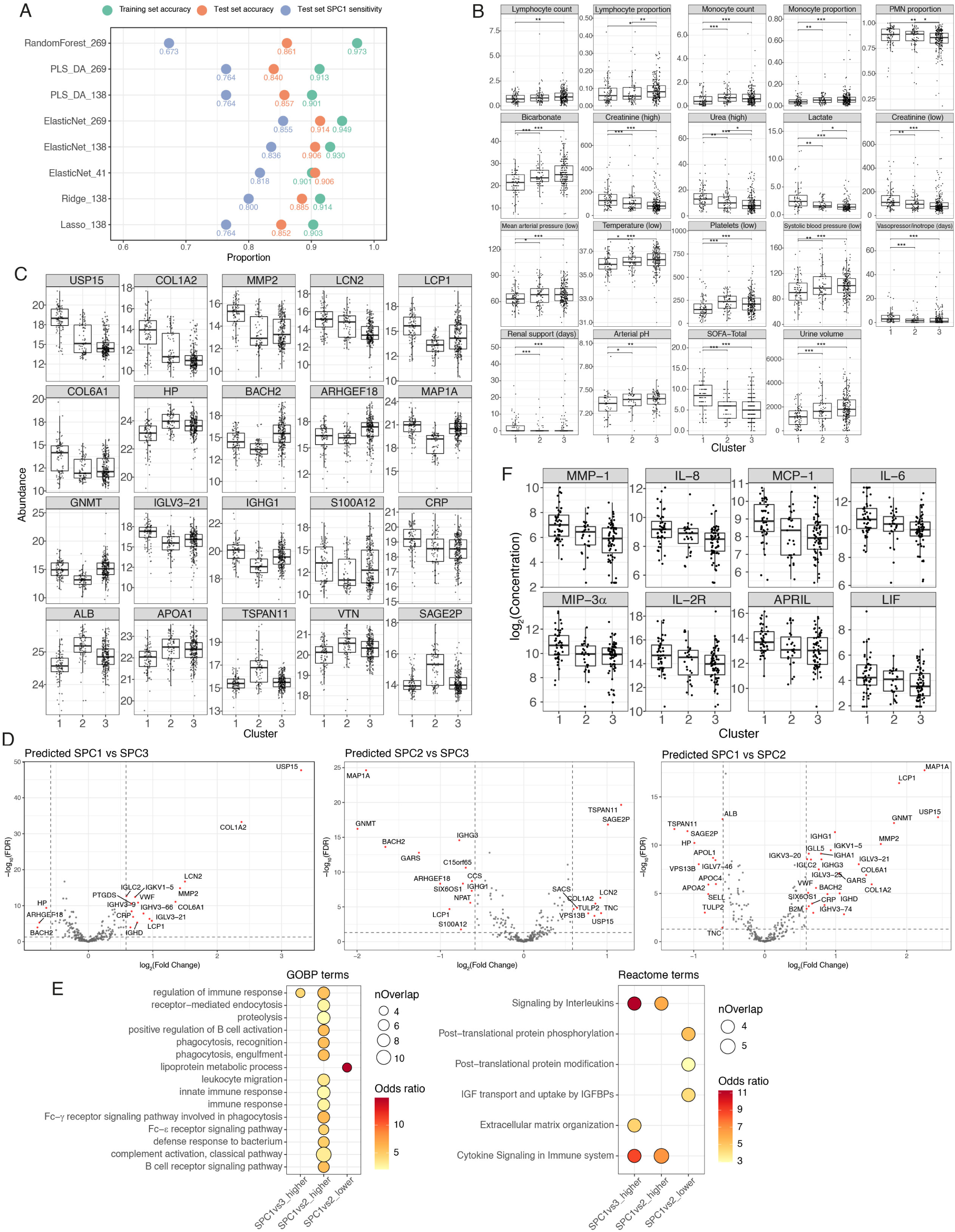
Validation and further characterization of the proteomic patient subgroups, related to Figure 5. (A) Performances of three-cluster SPC prediction models. Partial least squares discriminative analysis (PLS-DA), generalized linear models (GLM) and random forest (RF) class prediction models were tested using 80% of the sepsis discovery cohort samples for training (n=992) and 20% held-out samples for model evaluation (n=244). SPC assignments derived from consensus clustering in the sepsis discovery cohort were taken as the ground truth. Training and test set accuracy are based on three-cluster predictions. SPC1 sensitivity is the number of true SPC1 samples successfully predicted out of the total number of true SPC1. Numbers in model names indicate the numbers of protein candidates input to train each model. (B-E) Characterization of the validation cohort sepsis patient subgroups predicted from the best-performance model (“ElasticNet_269”). (B) Numerical clinical variables. (C) Representative protein abundance. (D) Comparison of protein profiles between the clusters. (E) Pathways enriched in the differentially abundant (FDR<0.05 and |FC|>1.5) proteins. Minimum overlap between data and annotation was set to 4. Only protein sets with any significant terms detected are shown. (F) Distribution of eight Luminex analytes differentially abundant between the clusters, in the subset of sepsis samples with Luminex data. Concentrations are in the unit of pg/mL.

**Figure S6.**
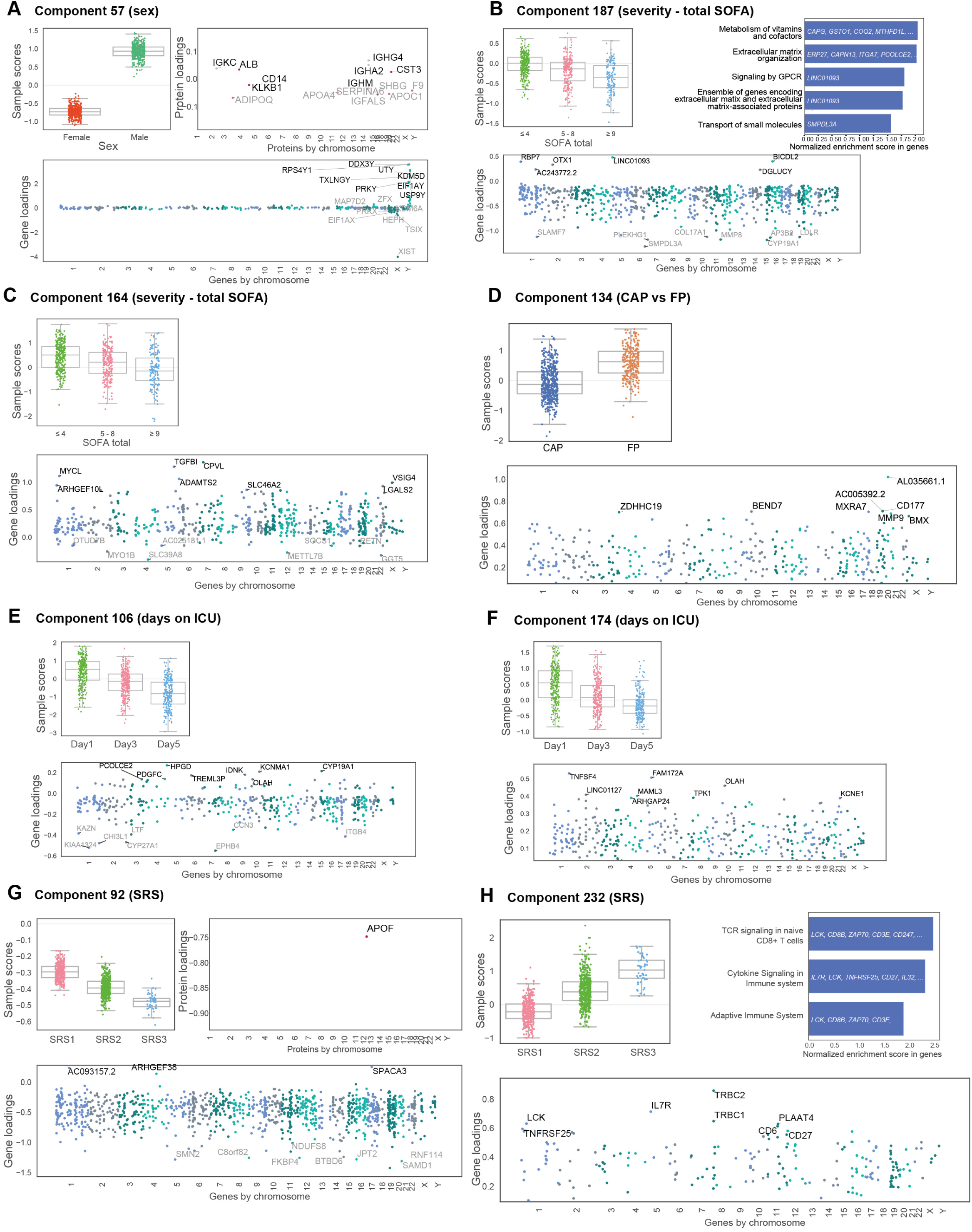
Integration with leukocyte transcriptomics, related to Figure 6. Matrix decomposition of proteomic and transcriptomic data for the same patients. Representative components are shown based on significance for association with clinical variables, SPC and SRS. For each component, the set of plots show sample scores by group (boxplots), protein or gene loadings for those significantly contributing to the component (posterior inclusion probability >0.5), z-scores of representative proteins (calculated within samples used for the matrix decomposition analysis), and pathway enrichment of significantly contributing genes. (A) Component most strongly associated with sex (component 57 *P_adj_*=1.3e-135, Mann-Whitney test). (B,C) The two most strongly associated components with total SOFA score, neither involving any plasma proteins (B) component 187, the component most strongly associated with total SOFA score (*P_adj_*=1.7e-18, rho=-0.35 Spearman) linking differential gene expression including nucleoside triphosphate catabolism enzyme *SMPDL3A*, aromatase *CYP19A1* and neutrophil collagenase *MMP8* (linked to higher SOFA scores) and showing overall pathway enrichment for ECM, vitamin and cofactor metabolism (C) component 164, the component second most strongly associated with total SOFA score (*P_adj_*=4.3e- 16, rho=-0.33) linking transforming growth factor-beta-induced protein *TGFBI*, probable serine carboxypeptidase *CPVL* and DNA-binding protein *MYCL* with lower SOFA score. (D) Component most strongly associated with source of sepsis (CAP vs FP) (component 134 P*_adj_*=1.9e-50, Mann-Whitney test) was restricted to gene expression. (E,F) Components most strongly associated with time of sampling following ICU admission, (E) component 106 P*_adj_*=1.0e-61 rho=-0.54 Spearman (F) component 174 P*_adj_*=1.5e-43 rho=-0.35. (G,H) Components most strongly associated with sepsis response signature (SRS) (G) component 92 P*_adj_*=1.6e-172, rho=0.78 Spearman (H) component 232 P*_adj_*=5.5e-168, rho=-0.78.

**Figure S7.**
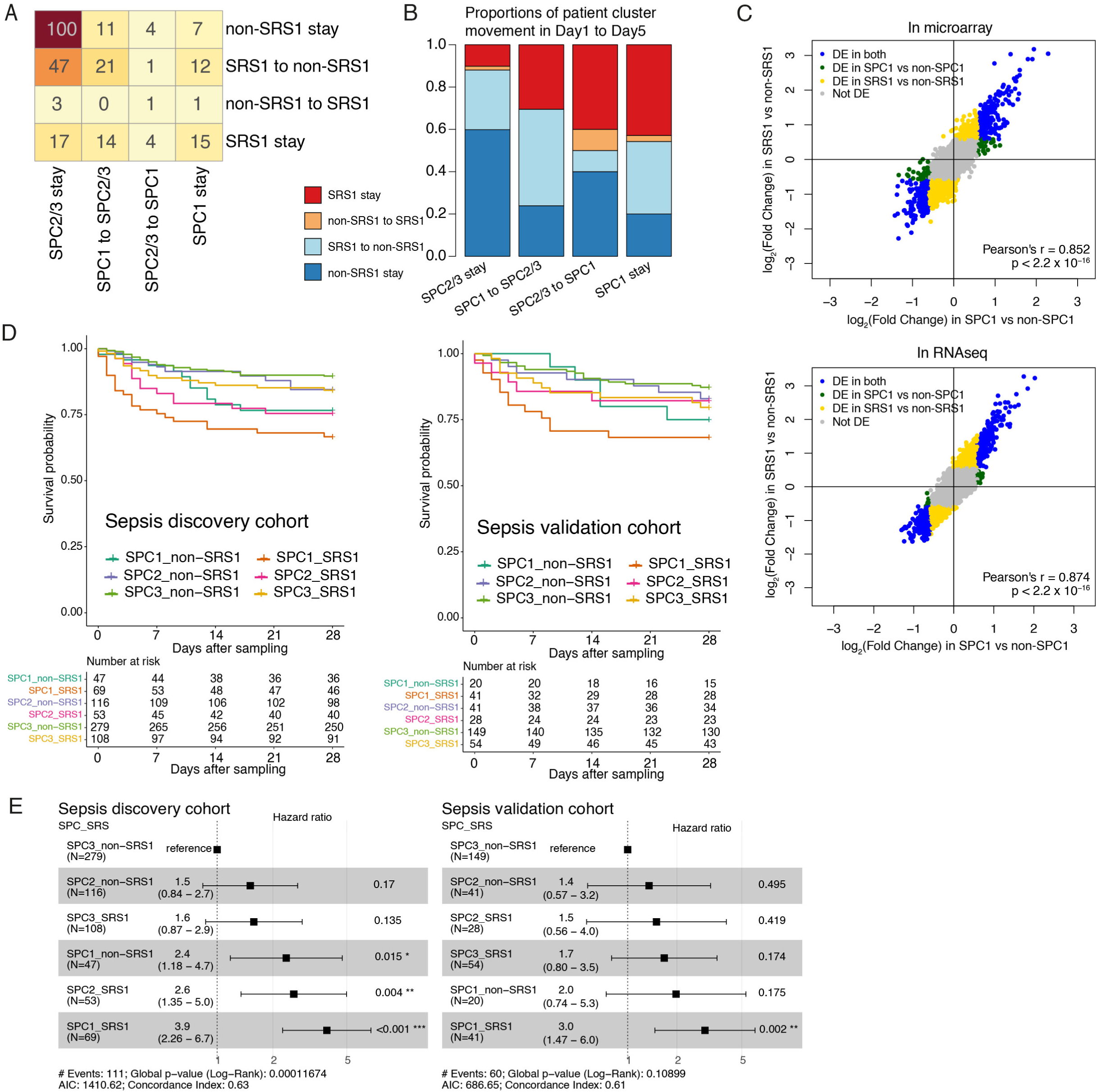
Interaction of the proteomic (SPC) and transcriptomic (SRS) patient subgroups, related to Figure 7. (A-B) Overlap between SRS and SPC cluster movement. Cluster movement observed in patients with at least two timepoints were used for comparison (n=258). Patients with 3 timepoints who moved in different directions over time were excluded. (A) Contingency table. (B) Stacked bar plots with heights of the bars corresponding to proportions of each SRS movement group in each SPC movement group. (C) Correlation plots for fold changes in comparing gene expression between SRS or SPC clusters. 1354 samples for 1010 patients had leukocyte gene expression data from either microarray or RNAseq. DE: differentially expressed (FDR<0.05 and |FC|>1.5). (D) Kaplan-Meier curves comparing survival probabilities at 28-day post-sampling between six groups of patients with a combined SPC and SRS classification. (E) Univariate Cox proportional hazard regression of 28-day mortality in six groups combining SPC and SRS classifications. Cluster assignments of the last available samples (within ICU Day 1/3/5) per patient with both SRS and SPC assigned were used. Hazards at 28-day post-sampling were compared.

## STAR Methods

### RESOURCE AVAILABILITY

#### Lead Contact

Further information and requests for resources and reagents should be directed to and will be fulfilled by Julian Knight.

#### Materials Availability

This study did not generate new unique reagents.

#### Data and Code Availability

The raw and processed timsTOF mass spectrometry data will be publicly available on the Proteomics Identification Database (PRIDE) (deposition in progress, accession ID TBC).

RNAseq gene expression data for GAinS study samples will be made available in the European Genome-Phenome Archive before publication. Microarray gene expression data for GAinS study samples are publicly available in ArrayExpress (E-MTAB-4421, E-MTAB-4451, E-MTAB-5273, and E-MTAB-5274).

### EXPERIMENTAL MODEL AND SUBJECT DETAILS

#### Cohorts

##### Abbreviation table

| Short abbreviation | HV (disc) | PreOp (disc) | PostOp (disc) | NoInf_ICU (disc) | Sepsis_ICU (disc) |
| --- | --- | --- | --- | --- | --- |
| Long abbreviation | Healthy (disc) | PreOp (disc) | PostOp (disc) | No infection ICU (disc) | Sepsis (CAP/FP) ICU (disc) |
| Full | Healthy volunteers (discovery cohort) | Pre-operative elective surgery cases(X-MINS, discovery cohort) | Post-operative elective surgery cases(X-MINS, discovery cohort) | No infection ICU cases (MOTION, discovery cohort) | Sepsis (CAP/FP) ICU cases (UK GAINs, discovery cohort) |
| Color scheme | Blue | Purple | Brown | Orange | Red |

| Short abbreviation | HV (vali) | PreOp (vali) | PostOp (vali) | NoInf_ICU (vali) | Sepsis_ICU (vali) | SepsisS_ICU |
| --- | --- | --- | --- | --- | --- | --- |
| Long abbreviation | Healthy (vali) | PreOp (vali) | PostOp (vali) | No infection ICU (vali) | Sepsis (CAP/FP) ICU (vali) | Septic Shock ICU |
| Full | Healthy volunteers | Pre-operative elective | Post-operative elective | No infection ICU cases (TACE and | Sepsis (CAP/FP) ICU cases | Septic Shock ICU cases (VANISH) |
|  | (validation cohort) | surgery cases (BIONIC, validation cohort) | surgery cases (BIONIC, validation cohort) | MONOGRAM, validation cohort) | (UK GAINs, validation cohort) |  |
| Color scheme | Blue | Purple | Brown | Orange | Red | Gray |

##### The UK Genomic Advances in Sepsis (GAinS) Study

###### Design

The Genomic Advances in Sepsis (GAinS) Study was an observational study of patients admitted to adult ICU with sepsis due to community acquired pneumonia or fecal peritonitis, as previously described (Davenport et al., 2016; Rautanen et al., 2015). Adult patients (>18y) were recruited at 34 UK ICUs between 2006 and 2019. Ethics approval was granted nationally (REC Reference Number 05/MRE00/38 and 08/H0505/78) and for individual participating centers, with informed consent obtained from patients or their legal representative.

In the GAinS study, sepsis was diagnosed according to the ACCP/SCCM guidelines (Sepsis-2) (Levy et al., 2003) as infection with signs of systemic inflammation with all patients showing organ dysfunction during ICU admission. Community acquired pneumonia (CAP) was defined as a febrile illness associated with a cough, sputum production, breathlessness, leukocytosis and radiological features of pneumonia which was acquired in the community or within two days of ICU admission (Angus et al., 2002; Walden et al., 2014). Fecal peritonitis (FP) was diagnosed at laparotomy as inflammation of the peritoneal membrane secondary to large bowel perforation and fecal contamination (Tridente et al., 2014).

###### Exclusion Criteria

Patient or legal representative unwilling or unable to give consent; age <18 years; pregnancy; an advanced directive to withhold or withdraw life sustaining treatment; admission for palliative care only; or immune-compromise.

###### Blood Sampling

Blood samples were collected on the first, third, and fifth day of ICU admission where possible. For the plasma samples, blood was collected into one 5ml EDTA vacutainer. The tube was inverted gently 10 times, and then centrifuged at 1600RCF for 10min at 4◦C. Four 500μL aliquots of the plasma layer was then transferred into cryotubes and stored at −20◦C until being transferred to Oxford in batches and stored at −80◦C.

###### Clinical phenotyping

Demographics and clinical measurements were recorded using an electronic case report form. Some of the clinical measurements including total white cell counts, blood pressure, heart rate, blood creatinine and bilirubin were recorded as the highest and lowest value measured on each day. Outcome in terms of death or survival was followed up for 6 months following ICU admission.

##### Other cohorts

Vasopressin vs Norepinephrine as Initial Therapy in Septic Shock (VANISH; REC reference 12/SC/0014) was a randomized clinical trial enrolling adult patients who had septic shock requiring vasopressors despite fluid resuscitation. Details of the study design and patient recruitment have been previously described (Gordon et al., 2016). The study reported no significant difference in outcome between vasopressin and norepinephrine groups, or between hydrocortisone and placebo groups. Up to four timepoints were included for each of the 45 VANISH patients in this study. The baseline timepoint (TP0) was mostly taken within 6 hours after the onset of shock and before the study drugs were given. Time from diagnosis of shock to study drug1 administration was within 6hr for 43 of the patients, and had a median of 4hr. Blood samples were collected into 10mL EDTA vacutainers on ice. The tube was inverted 5-8 times and then centrifuged at 1000RCF for 10min before the plasma layer was aliquoted.

The Oxford BioBank (OBB; REC reference 18/SC/0588) is a random, population-based biobank of healthy participants initially recruited between the ages of 30 and 50 years from the Oxfordshire general population (Karpe et al., 2018). Individuals with: previous diagnosis of myocardial infarction or heart failure currently on treatment; untreated malignancies; or other systemic ongoing disease, and pregnant women were excluded. One sample was included for each of the 152 OBB participants in this study. To match the demographics of other cohorts as closely as possible, older OBB participants were selected. Plasma samples were collected with EDTA as the anti-coagulant.

X-MINS (Acquired loss of cardiac vagal activity is associated with myocardial injury in patients undergoing non-cardiac surgery; REC reference 16/LO/0635) was an observational mechanistic cohort study at the University College London Hospital, recruiting adult patients undergoing major elective noncardiac surgeries including orthopedic, upper GI and colorectal surgeries. The X-MINS cohort has been utilized to examine whether serial measures of cardiac vagal dysfunction were associated with perioperative myocardial injury and noncardiac morbidity (May et al., 2019), and had measured microRNA concentrations in extracellular vesicles (May et al., 2020). Two samples for each of the 106 X-MINS patients were included in this study, one taken pre-operation (PreOp (disc)) and one taken within 24hr post-operation (PostOp (disc)). Blood samples were collected into EDTA vacutainers and centrifuged at 3500RCF for 10min before the plasma layer was aliquoted sterile filtered.

BIONIC (Biomarker based Identification Of Nosocomial Infective Complications; REC reference 14/EM/1223) was a prospective observational study at the Royal London Hospital that recruited patients undergoing elective major abdominal surgeries encompassing colorectal, upper GI (gastrointestinal) and HPB (Hepato-Pancreato-Biliary) surgeries. For each of the 43 patients included in this study, three samples were included, taken either immediately before induction of anesthesia (PreOp (vali)), or 24hr post-operation (PostOp (vali)), or 48hr post-operation. Ten patients also had a 2-6hr post-operation sample included. Blood samples were collected into 4.5mL citrate vacutainers and centrifuged at 1860RCF for 10min before the plasma layer was aliquoted.

The MOTION (Methylnaltrexone for the Treatment of Opioid Induced Constipation and Gastrointestinal Stasis in Intensive Care Patients; REC reference 14/LO/2004) trial recruited adult ICU patients who were mechanically ventilated, receiving opioids and were constipated (Patel et al., 2020). For the 50 patients included in this study, reasons for ICU admission included non-operative medical causes (n=34) and emergency(n=13) or elective (n=3) operative procedures. Patients admitted to ICU for infection or respiratory causes were not included. One sample from each patient, taken before the study drug was given, was included for this study. Blood samples were collected into citrate vacutainers on ice and centrifuged at 1500RCF for 15min before the plasma layer was aliquoted.

The MONOGRAM (REC reference 15/LO/0933) study was set up for diagnosing and monitoring infection in critically ill patients using metabolic and immunological signatures. Baseline samples taken within 48hr of ICU admission for 14 mechanically ventilated patients with systemic inflammatory response syndrome (SIRS) but without identified infection were included in this study. Blood samples were collected into 10mL EDTA vacutainers on ice. The tube was inverted 5-8 times and then centrifuged at 1000RCF for 10min before the plasma layer was aliquoted.

TACE (REC reference 10/H0709/77) is a general abbreviation for observational studies on the mechanisms of monocyte priming and tolerance in vitro and in vivo to ascertain TNF-α converting enzyme (TACE) activity and metabolic signatures of patients with direct and indirect acute lung injury (ALI). Adult patients with or at risk of ALI, admitted to ICU within Imperial College Healthcare

NHS Trust, were recruited within 48hr of onset of ALI or intubation. More details were described in associated publications (Antcliffe et al., 2017, 2018; O’Callaghan et al., 2015). The baseline samples on study entry for twelve patients with noninfectious SIRS (ten with brain injury, one with cardiac arrest, one with motor neurone disease) were included in this study. Plasma samples were collected with EDTA or heparin as the anti-coagulant.

### METHOD DETAILS

#### TimsTOF mass spectrometry

##### Sample preparation of non-depleted plasma for high-throughput LC-MS/MS platform

Plasma samples were thawed at 4◦C. 50μL aliquots of each of the 2622 clinical plasma samples from the 8 cohorts were pipetted into 28 96-well plates in randomized positions. Three microliters of plasma were then diluted in 50mM ammonium bicarbonate at 1:30 ratio (sample: buffer) in 96-well plates. Plates were centrifuged at 2000g for five minutes before transferring thirty microliters of diluted plasma into clean 96-well plates for subsequent in-solution digest using the BravoAssaymap liquid handler robot (Agilent). In brief, samples were reduced with DTT (15mM final concentration) for 30 minutes at room temperature, alkylated with iodoacetamide (70mM final concentration) for 30 min at room temperature followed by the addition of 100 μl of 50mM ammonium bicarbonate containing 10mM DTT to quench the reaction. Samples were incubated with 1 μg of Trypsin (Worthington TPCK) at 37C overnight. Trypsin digestion was stopped by adding trifluoroacetic acid at 1% final concentration.

##### Pools

A pool was built using thirty microliters of the diluted plasma for each patient. This pool was included in each digestion plate as a “QC Plate pool” to monitor trypsin digestion. An additional pool was created post digestion (labelled as QC system) and run every twenty-four samples to monitor the performance of the LC system across the whole dataset.

##### Generation of a plasma library combining top 64 depletion with High pH fractionation

Plasma has a high dynamic range at protein level. To increase the detection range of proteins in plasma and to generate a protein library we created a super-depleted plasma by combining two-consecutive antibody-based depletions using first the SepproIgY14 column (SEP030 column Sigma-Aldrich) to deplete for the top 14 most abundant plasma proteins followed by a Seppro Supermix column (SPE050 column, Sigma-Aldrich). 200μL of the pooled plasma was diluted to 1ml with ‘Dilution buffer’ containing 0.1%N-Octyl-B-D-glucopyranoside then applied to a prewashed Costar 45uM spin filter and cleared by centrifugation 14,000 x g for 10mins at room temperature. 1ml of the filtrate was injected into a BioRad NGC medium pressure system following the method described (Keshishian et al., 2017), with some additional changes to accommodate the use of the two columns. In brief, the two depletion columns were attached in tandem, first passing the Seppro IgY14 column then the Supermix column resulting in the unbound depleted plasma directly flowing into the second column. The columns were pre-equilibrated for 3.33mins at 1.5ml/min in dilution buffer followed by sample application at 1ml/min for 1min then 0.5ml/min for a further 3mins. Flow rate was then increased to 1.5ml/min and dilution buffer isocratically for 15mins. The column was then stripped of the proteins which had been depleted, using stripping buffer at a flow rate 2mls/min for 6mins. Depleted proteins were collected in 0.5ml fractions between min 12.33 and 22.33 and in 1ml fractions until min 28.33. The column was then neutralized and re-equilibrated into dilution buffer for 9.33mins at 1.5ml/min. 50μL of each fraction was evaluated using Ag stain and the depleted fractions pooled together according to the Abs280nm. The plasma elution profile showed that the depleted peak appeared between min 3.5 and 13.5 and accounted for approximately 40% of the total protein present in the sample. The fractions were pooled into 15 pools containing approximately 3% total proteins present and labelled ‘A’ to ‘O’. Each fraction ‘A’ to ‘O’ was chloroform/methanol precipitated and the pellets resuspended into 50μL of 6M Urea in 0.1M Tris pH 7.8. To ensure protein pellets were solubilized, samples were mixed at room temperature for 20min and sonicated for 10min in a sonicating water bath. Urea concentration was diluted down to 1M by adding 250μL H_2_O. Samples were incubated with 1μg of smart trypsin (ThermoFisher) at 70°C for an 1h shaking at 300rpm. 150μL of the sample was then HpH fractionated into 12 fractions on RP-W cartridges using the Bravo Assaymap liquid handler (Agilent technologies). Each individual fraction was then loaded onto C18 EvoTips for subsequent LC-MS/MS analysis.

##### LC_MS/MS using the high-throughput Evosep One - Bruker timsTOF Pro platform

Samples were analyzed using an Evosep One LC system connected to the timsTOF Pro mass spectrometer (Bruker Daltonics) (COvid-19 Multi-omics Blood ATlas (COMBAT) Consortium, 2022). Briefly, Peptides were analyzed using the pre-built 100 samples / per day method (EvosepOne) with an 11.5 min gradient (total cycle time of 14.4min) at a 1.2 µl/min flow rat (Bache et al.). In brief, tryptic peptides were transferred from the pre-loaded C18 Evotips with a pre-built gradient to a sample loop and separated on an 8cm C18 analytical column (Evosep Pepsep, 3um beads, 100 um ID) with an overall gradient from 3 to 40% acetonitrile.

Mass spectrometry data were acquired in PASEF mode (oTOF control v6.0.0.12). The ion mobility window was set to 1/k0 start = 0.85 Vs/cm2 to 1/k0 end = 1.3 Vs/cm2, ramp time 100 ms with locked duty cycle, mass range 100 - 1700 m/z. MS/MS were acquired in 4 PASEF frames (3 cycles overlap). Target intensity was set to 6000 and threshold intensity 200. The data acquisition method has been deposited with the raw data in the PRIDE data repository (deposition in progress, accession ID TBC). To assess the technical reproducibility, QC system pools were run every twenty-four samples. QC plate pools were run to monitor the performance of the trypsin digestion in each plate.

##### QE-HF mass spectrometry

###### Sample preparation of top 12 depleted plasma analyzed using a low throughput platform (QE-HF)

Five microliters of blood plasma collected from acute inflammation patients were depleted for the 12 most abundant proteins using spin columns (Pierce, Waltham, MA). Depleted plasma protein solutions were then precipitated using TCA/DOC. Protein pellets were digested using Smart digest kit (ThermoFisher) and desalted on SOLA SPE cartridges as described in (Prianichnikov et al., 2020).

Peptides were injected into a LC-MS system comprised of a Dionex Ultimate 3000 nano LC and a Thermo Q-Exactive HF. Peptides were separated on a 50-cm-long EasySpray column (ES803; Thermo Fisher) with a 75-µm inner diameter and a 60-minute gradient of 2 to 35% acetonitrile in 0.1% formic acid and 5% DMSO at t flow rate of 250 nL/min. MS1 spectra were acquired with a resolution of 60,000 and AGC target of 3e6 ions for a maximum injection time of 45 ms. The Top12 most abundant peaks were fragmented after isolation with a mass window of 1.2 Th at a resolution of 30,000. Normalized collision energy was 28% (HCD).

##### Luminex assay

Sixty-five inflammatory mediators were measured in 204 samples from 146 patients from the GAinS study using the ProcartaPlex™ Luminex platform (ThermoFisher Scientific, Waltham, MA, USA), as has been described previously (Antcliffe et al., 2022). Briefly, plasma samples were measured across three 96-well plates with data acquired on Luminex 100 system. For each analyte, background fluorescence was corrected for and the lower/upper limits of quantification (LOQs) were determined. Absolute concentrations were derived from concentration-response standard curves fitted with a five-parameter logistic model for each analyte on each plate. Sample concentrations beyond the LOQs were censored at the LOQ values. Outlying samples of poor quality were identified and removed.

##### Leukocyte whole-genome expression profiling

For the GAinS patient-timepoints included in this study, 1354 (1010 patients) have paired leukocyte gene expression data available through either microarray profiling (518 patients, 642 samples), RNA sequencing (649 patients, 837 samples), or both platforms.

The microarray datasets were acquired using Illumina Human-HT-12 v4 Expression BeadChips, full details of which have been described previously (Davenport et al. 2016; Burnham et al. 2017). Four batches of microarray data have been cleaned-up, combined and the batches normalized. Gene expression was represented at gene-level for 22130 genes. The RNAseq dataset was generated on NovaSeq with 100bp paired end reads across 12 lanes. After quality control, gene expression was represented as TMM (trimmed mean of M-value)-normalized log-transformed counts per million for 20416 features.

The SRS transcriptomic endotypes have been assigned based on these datasets as well as for 7 samples with qPCR data using a 7-gene set random forest model previously described (Cano-Gamez et al., 2022). For the small number (11) of border-line samples that had different assignments from the platforms, assignments were taken in the order of priority as microarray before RNAseq before qPCR. Since there was only a small number (48) of patient-timepoints assigned to SRS3 (transcriptionally closer to healthy volunteers) and that SRS3 patients are more similar to SRS2, SRS3 was combined with SRS2 as a ”non-SRS1” category in the analysis of this study.

### QUANTIFICATION AND STATISTICAL ANALYSIS

#### TimsTOF protein identification and quantification

Data was analyzed by the FragPipe pipeline consisting of FragPipe 14.1, MSFragger 3.1 (Kong et al., 2017; Yu et al., 2020), Philosopher 3.3.11 (da Veiga Leprevost et al., 2020) and Python 3. Blank runs were excluded and each file defined as Experiment to facilitate LFQ. Data was searched against a fused target/decoy database generated by Philosopher and consisting of human UniProt SwissProt sequences, plus common contaminants. The database had 40860 entries (including 50% decoy entries). MSFragger parameters were set to allow a precursor mass tolerance of plus/minus 10ppm and a fragment tolerance of 20ppm. Isotope error was left at 0/1/2 and masses were set to re-calibrate. Protein digestion was set to semi specific trypsin with up to 2 allowed missed cleavage sites, allowing peptides between 7 and 50 residues and mass range 500 to 5000 Da. N-terminal protein acetylation and Methionine oxidation were set as variable modifications. ID validation was done with PeptideProphet (Keller et al., 2002) and ProteinProphet (Nesvizhskii et al., 2003) with default settings.

Label free quantitation was conducted with IonQuant (Yu et al., 2021) and Match-Between-Runs enabled with 10 selected pool injections from the beginning and end of the sample queue (tag “lib” in experimental design tab) in addition to the 10 best correlating runs for each file, using Top-3 quantitation. Feature detection tolerance was set to 10ppm and RT Window to 0.6 minutes with an IM Window of 0.051/k0. For matching, ion, peptide and protein FDRs were relaxed to 0.1. MBR top runs was set to 10 in addition to selected runs tagged “lib”.

#### TimsTOF data pre-processing

From the FragPipe output of label-free quantifications (LFQ) of 4570 sample and quality control injections, we subsetted and used razor intensities of 2782 proteins identified by at least one unique peptide. This was a sparse matrix since the majority of proteins were only detected in the fractionated libraries instead of in actual samples. Zeroes were taken as missing values. Gene names for the proteins were retrieved by mapping accession IDs on the UniProt protein knowledgebase (Bateman et al., 2021).

##### Protein and sample filtering

In order to retain proteins that reach a detectable level in only certain biological conditions, protein detection was counted within each of the biological groups. Among the cohorts, the GAinS study is the majority group constituting >70% samples. Filtering based on the heterogeneous syndrome of sepsis as one group could potentially miss out the proteins with distinct patterns between unknown biological subgroups. Therefore, the GAinS samples were further separated by unsupervised consensus clustering on a matrix of binary states detected (1) or missing (0), which identified four GAinS protein detection subgroups, the smallest of which comprised 292 samples. Protein detection was then counted within nine groups including: the healthy volunteers of OBB; the pre-operation samples of X-MINS and BIONIC; the post-operation samples of X-MINS and BIONIC; the no-infection ICU samples of MOTION, MONOGRAM and TACE; the septic shock samples of VANISH; and the four GAinS detection groups (Figure S1B). We kept 291 proteins that were detected in at least 50% sample injections in at least one of the nine groups.

Sample quality represented by sample-wise protein detection numbers showed variation corresponding to the changes of the chromatographic column. We excluded 32 sample injections for having outlying low detections (<125 proteins), the threshold of which was determined by plotting the detection numbers (Figure S1C). The raw intensity data matrix post-filtering contained quantifications for 291 proteins in 2615 sample injections.

##### Correction for cell residue contaminations

After filtering, the data was normalized, missing values imputed and batches corrected to derive an “interim processed data” version, as shown in the flowchart (Figure S1A). Although all plasma samples were extracted by the standard approach of centrifuging whole blood taken with anticoagulants, samples from different studies recruited in a variety of clinical settings could potentially have differences in pre-analytical handling. Even within a single study following the same protocol, in the interim processed data the GAinS samples, recruited over 14 years in 34 ICUs, showed a small subset of ∼12% samples that were significantly higher in proteins including actins and actin-binding proteins that point to higher platelet residues in plasma. Therefore, it is necessary to assess and account for the impact of the technical effects.

We calculated three contamination indices (Figure S1D) to assess plasma sample contamination by erythrocyte lysis, platelet contamination, or partial coagulation, based on the protein marker panels proposed by Geyer et al. (Geyer et al., 2019). We calculated and considered the contamination indices using both the log-transformed raw intensities and the interim processed data as both have uncorrected biases. X-MINS samples were collected using the strictest protocol (higher speed in centrifugation and filtered through 0.8μm membrane filters) and were among the samples with the lowest level and least variation in the cell residues (erythrocytes index and platelets index) (Figure S1D). As the coagulation index was less variable across the cohorts, and as sepsis pathophysiology involves dysregulated thrombosis, the difference in coagulation index was considered to be biological signal and not corrected for.

We corrected for cell residue contamination by identifying and removing the most affected proteins species from the dataset. In addition to proteins on the marker panels, we also removed proteins that correlated with either the platelet or erythrocyte index in both the raw and interim processed data (Pearson’s r > 0.20). In total 22 proteins were removed, leaving 269 to be further processed (Figure S1E). The potentially contaminated samples did not need to be excluded, since these were not outlying in protein detection numbers or on PCA after excluding the contamination proteins, and the removed proteins did not compromise the detection of other proteins.

##### Normalization and imputation

We used Variance Stabilizing Normalization (VSN) (Huber et al., 2002) to normalize the raw intensities and account for systematic bias. This method was selected based on running a smaller sepsis pilot dataset through Normalyzer (Chawade et al., 2014) which compared different normalization algorithms, as well as based on others’ work showing the advantage of VSN in addressing the variance heterogeneity in mass spectrometry-based proteomics (Karp et al., 2010; Välikangas et al., 2016).

Assuming proteins are missing not completely at random, we applied a hybrid imputation approach. For 170 proteins detected in ≥60% of the samples, we applied the K-Nearest Neighbors (KNN) algorithm which is more tolerant on the assumption of the causes of missingness. For each protein with missing values to be imputed, KNN finds the k (k=6) most similar proteins (neighbors) based on the non-missing values, and fills in the missing value by the mean observed for the neighbor proteins in this sample (Hastie et al., 2021). When there is a higher proportion of missingness in a protein, KNN is not suitable since neighbor proteins cannot be accurately located. For the 99 proteins detected in <60% of the samples, missing values were imputed by random draw from a protein-specific normal distribution with a standard deviation (s.d.) of 30% s.d. of the measured values and a down-shift of the mean by 1.8 s.d.

##### Batch correction

The 28 randomized sample plates were profiled in a single mass spectrometry experiment but the long acquisition duration introduced technical variation. We performed batch correction across the plates using the ComBat function from the sva package (Leek et al., 2021). Seventeen batches were determined based on the pattern of PC1 scores of the plates in uncorrected data and the spectral yields and detection numbers in spiked-in pool injections repeated along the plates. Seven plates that were the most consistent between each other were used as the reference batch where measurements were not altered.

##### Discovery and validation cohorts

For downstream analysis, we defined the discovery and validation cohort sets from the sepsis and control cohorts (Figure 1A). The GAinS and OBB patients were each separated into a discovery and a validation cohort with a split of 2:1 by random draws after pre-processing. Serial samples from the same patient were kept together.

#### QE-HF data pre-processing

Mass spectrometry data were analyzed using Progenesis QI software combined with PEAKS v8.5 as a data search engine using standard settings. 1356 proteins were identified by at least one unique peptide in the raw intensity output. The 0.5% lowest measurements were considered to be machine noise and removed from the distribution. Proteins missing in more than 30% samples were filtered out, leaving 1123 proteins in the cleaned-up dataset. All samples passed quality control. The dataset was then normalized by VSN and imputed by KNN.

#### Pathway and network analysis

Gene names were mapped from the UniProt accessions using the UniProt (Bateman et al., 2021) retrieve/ID mapping tool. Pathway enrichment analysis was performed using the XGR R package (Fang et al., 2016) with annotations from Gene Ontology Biological Process (GOBP), Gene Ontology Cellular Component (GOCC), or the Reactome pathway database. Annotation items with a size between 5 and 2000 and a minimum overlap with input data of 5 were included for the enrichment tests, unless otherwise specified. Significantly enriched terms were defined by FDR < 0.05 in hypergeometric tests with all proteins or genes detected in the experiment (including in fractionated and depleted library samples for proteins) as the background. Pathway enrichment in proteins differentially abundant between SPC (Figure 4G, S5E) were tested in proteins with higher or lower abundance in each contrast separately, with minimum overlap between data and annotation set to 4.

Protein-protein interaction (PPI) data was retrieved from the STRING v11 database with a confidence score cut-off of 0.7 and zero additional interactors. The PPI network was constructed and visualized through the Cytoscape v3.8.2 platform (Shannon et al., 2003) using perfuse force directed layout. The main network comprising 141 proteins was then divided into clusters by the Markov cluster algorithm applied in the “clusterMaker” plugin (Morris et al., 2011). Proteins in each clusters were tested for GOBP enrichment, using a minimum overlap of three for the six smaller clusters. Node color was mapped to Pearson’s correlation coefficients of PC1 scores and the protein level across samples, as higher PC1 score was shown to correlate with higher disease severity towards the sepsis end in Figure 1B.

The protein co-expression network was constructed using the WGCNA R package (Langfelder and Horvath, 2008) and visualized using Cytoscape. The soft thresholding power of 3 was chosen based on the lowest power for which the scale-free topology fit index curve reaches a high value (>0.85). For network construction we used a signed hybrid type of network, unsigned type of topological overlap matrix (TOM), biweight mid-correlations, and a maximum 5% of samples that can be considered as outliers on either side of the median. The minimum module size was set to 5 considering the smaller number of features in proteomics compared with transcriptomics datasets. For visualization of the co-expression network (Figure 1F), nodes with larger width and height have higher intramodular connectivity, which is the sum of adjacency with all other nodes in the same module. Exact positions of the nodes have been manually adjusted to display all the labels. Only edges with topological overlap ⩾0.02 are included thus 180 out of the 184 nodes with a co-expression module assigned are shown. For visualization of the topological overlap matrix (TOM) (Figure S1G), dissimilarity (1-TOM) was raised to the power of 15 to minimize effects of noise and spurious associations. Distance used in the dendrogram is based on the topological overlap between two proteins nodes, which is a function of the adjacency between the two nodes and between either node with all other nodes. Correlation of module eigengenes with clinical characteristics and dummy-coded sepsis-control contrasts were calculated using Spearman’s correlations, shown in Figure 3C.

In the single sample gene set enrichment analysis (ssGSEA), the enrichment scores of gene sets were calculated based on walking down a list of proteins ranked by their intensity in each single sample. Through the ssGSEA projection the matrix of protein intensities in samples is converted to a matrix of enrichment scores of gene sets in samples. This projection was performed using the Gene Pattern platform (Reich et al., 2006). Gene sets with less than 10 overlaps with the input proteins were excluded, identifying 296 gene sets using the GOBP annotation. PCA on the projected matrix revealed the gene sets that differentiated among the samples. Eight gene sets with top loadings on PC1 and 8 gene sets with top loadings on PC2 were labelled (Figure 1D).

#### Bioinformatic and statistical analysis

##### Exploratory analysis

We performed principal component analysis on all samples in the pre-processed data using the prcomp R function without scaling. We explored the relationship between the first four PCs or individual proteins with sepsis severity by testing Spearman correlation with the clinical variables, restricted to the first available samples of GAinS patients. Clinical variables detected in less than 30% of patients were excluded. Variables only available in certain groups of the patients (e.g. estimated days from CAP/FP onset) were retained. All statistical analysis were performed in R (R Core Team, 2015).

##### Statistical tests

Differential abundance analysis for proteins and differential expression analysis for genes were performed by fitting the intensities in linear models using the limma R package (Ritchie et al., 2015), using only the first available sample of each patient and including age and sex as covariates. The Benjamini-Hochberg procedure was used to adjust for multiple testing. Significance for downstream analysis was defined as FDR < 0.05 and fold change (FC) > 1.5 unless otherwise specified. Comparisons between the pre- and post- operative samples are paired and with no additional covariates. All tests are two-sided.

Welch’s t-test, also known as unequal variances t-test, is more conservative when the group size and variance differs by a large amount between the two groups. The differential abundance analysis between GAinS and MONOGRAM+TACE was also performed using Welch’s t-test and highly consistent results were obtained as when using limma (Pearson’s r for log_2_FC > 0.99).

Cytokine concentrations measured by Luminex assay were compared by Wilcoxon rank-sum tests (i.e. Mann-Whitney tests) using the first available sample of each patient.

Categorical clinical variables were compared using Chi-squared tests without Yate’s correction. Numerical clinical variables were compared between two groups using Wilcoxon rank-sum tests. For comparing numerical clinical variables between three groups, Kruskal–Wallis test (i.e. one-away analysis of variance on ranks) was used to determine whether there was any significant difference (FDR<0.05) between the groups. For variables where the null hypothesis in Kruskal–Wallis tests were rejected, Dunn’s post-hoc tests were performed to compare between each pair of the three groups, the significance levels of which were labelled on boxplots after adjusting for multiple testing within the variable. SOFA scores and ARDS levels were considered as numerical variables. Only the first available samples following ICU admission of each patients were included in comparing the total of 66 clinical variables.

Survival differences were assessed and Kaplan-Meier curves plotted using the R packages survival and survminer (Kassambara et al., 2021; Therneau and Grambsch, 2000). The input data is a data frame specifying the time to event from the sampling day, the event (death or end of 28-day or 6-month observation) and the patient subgroups. For patients with multiple timepoints sampled, cluster assignment from the last available sample within five days of ICU admission was used. The p values were given by log-rank tests. Hazard ratios and the confidence intervals were calculated using univariate (SPC) or multivariate (SPC and SRS) Cox proportional hazard models.

##### Data visualization

On all boxplots, hinges show the first and third quartiles on both sides of the median, and the whiskers extend to the highest or lowest values within 1.5*IQR of the hinges. Data points are plotted on horizontally jittered positions on the boxplots. For sepsis patients with serial sample points, only the first available samples following ICU admission were plotted.

On all volcano plots, proteins more abundant in Group A of the contrast “Group A vs B” are plotted on the right-hand side of the plot. Differentially abundant proteins (FDR<0.05 and |FC|>1.5) are plotted in red points.

Trajectories of patients’ movements between the proteomic clusters were visualized using the ggalluvial package (Brunson, 2020), restricted to patients with a baseline (Day1) sample available. Widths of the flows on the y axis are scaled to the number of patients in each movement type. Figure 5F and a subset of the icons in Figure 1A “Downstream analysis” panel were generated with BioRender.com.

##### Unsupervised clustering

We applied consensus clustering to generate unsupervised clusters in 1236 discovery cohort GAinS samples, using the ConsensusClusterPlus R package (Wilkerson and Hayes, 2010). Compared with a single-run hierarchical clustering performed on all available samples and features, the consensus clustering approach has the advantage of cluster results being robust to small variations in sample or feature composition. For each cluster number k tested from two to ten, 500 iterations were performed, randomly sampling 80% of samples and 90% of proteins in each iteration. In each iteration, unsupervised hierarchical clustering was performed based on Euclidean distance for dissimilarity between samples and Ward’s method as linkage for cluster agglomeration.

The cumulative distribution function (CDF) curves (Figure 4C) were inspected to assess cluster stability along an increasing number of clusters (k). The increases in area under the CDF were the largest when k increased from 2 to 3, and reached an elbow point at k=4 (Figure 4D). We also assessed how model accuracy improved as cluster number increased by calculating the total within-cluster variation in 10-fold cross-validation with k-means clustering. The fit of test points to the nearest centers improved gradually as the cluster numbers increased from 1 to 10, and the step widths were the largest from k=1 to k=3. Therefore, k=3 was chosen as the optimal number of partitioning for this dataset, balancing the need for explaining most of the variability and for the simplicity of describing the major patient subgroups.

##### Cluster prediction models

To validate the three patient clusters (SPC) in independent samples, cluster prediction models were built based on SPC assignments in GAinS discovery cohort samples, and the model with the highest accuracy was applied to the GAinS validation cohort samples. In evaluating the prediction model performances, accuracy is the sum of the diagonal of the contingency table divided by the total number. A random 3-cluster classification keeping the same proportions in each cluster as in the discovery cohort would have an expected accuracy of (21%)^2^+(24%)^2^+(55%)^2^ = 40%. Sensitivity for a particular event is the proportion of events correctly predicted, out of the number of true events. The best-performing model was selected based on the test set accuracy.

We tested eight models in total for SPC prediction using three statistical learning methods: partial least squares discriminative analysis (PLS-DA), generalized linear models (GLM), and class prediction by random forest through the R packages ropls, glmnet, caret and randomForest (Friedman et al., 2010; Kuhn, 2021; Liaw and Wiener, 2002; Thévenot et al., 2015). Input candidates are either the total 269 proteins in pre-processed data, or restricted to the 138 proteins that were differentially abundant between any pair of the 3 discovery cohort clusters at a less stringent threshold (FDR<0.05 and |FC|>1.2), or the 41 proteins at a larger fold change (FDR<0.05 and |FC|>1.5).

The numbers of predictive components to include in PLS-DA models were determined by calculating the proportion of variation explained in 7-fold cross-validation and evaluating model significance with label permutations GLM models were tested with lasso, ridge, and elastic net regressions to include the overfitting penalty and enable variable selection. Symmetric multinomial models were fit so that each of the 3 SPC classes were represented by a GLM. Two parameters were optimized: the tuning parameter λ which controls the overall strength of the penalty by defining the amount of coefficient shrinkage; and the elastic net mixing parameter α which controls whether the penalty behaves more towards a lasso (α=1) or ridge (α=0) regression for correlated predictors.

For lasso and ridge regressions, the value of λ was determined by minimizing the prediction error rate in 10-fold cross validation (CV) within the training set. When there was not a great increase in CV misclassification rate between the λ that gives the smallest error, and the largest λ such that the error is within the smallest error plus one standard error, the latter value of λ was chosen so that the model was less heavily based on the training set. For elastic net regression (0<α<1), the best values for λ and α were selected by testing a grid of 20*20 combinations, and selecting the combination that gave the highest prediction accuracy in cross validation.

Since the elastic net model with 269 protein candidates had the best performance in GLM models, 269 proteins were used as candidate inputs to train the random forest model. The 3-cluster accuracy from 10-fold cross validation was used as the optimizing metric for tuning the parameters. In the optimized random forest model, 35 variables were randomly sampled for each iteration; the maximum number of terminal nodes of each tree was 42; 350 trees were trained; the other parameters were kept as default values in the package.

The 8 protein predictors in the minimal elastic net model were selected by restricting to 3 proteins with the strongest signals in the differential abundance analyses between each cluster pair, whilst also ensuring at least one higher- and one lower- abundance protein were retained for each pairwise contrast.

##### Matrix decomposition

We performed matrix decomposition on the 837 samples with both RNAseq (for 20,416 genes) and plasma proteome data (for 269 proteins), using the SDA method (Hore et al., 2016), with the same number of iterations (10), clustering and cutoffs to end up with 284 latent components. Within the components, we also used the same cutoff for the posterior inclusion probability (0.5) as (Hore et al., 2016) for the inclusion of the gene or protein loading scores. This resulted in 76 components with protein loading scores. Similarly to our work in (COvid-19 Multi-omics Blood ATlas (COMBAT) Consortium, 2022) we matched these to clinical features as well as the SRS or SPC using either Spearman’s correlation coefficient or Mann-Whitney U rank test, adjusting for multiple testing using the Benjamini/Hochberg (non-negative) method. Correlation of the latent components with SRS were tested with the quantitative SRS scores (Cano-Gamez et al., 2022). Z-scores of representative proteins shown in Figure 6 were calculated within the first available samples used in matrix decomposition analysis.

### ADDITIONAL RESOURCES

#### KEY RESOURCES TABLE

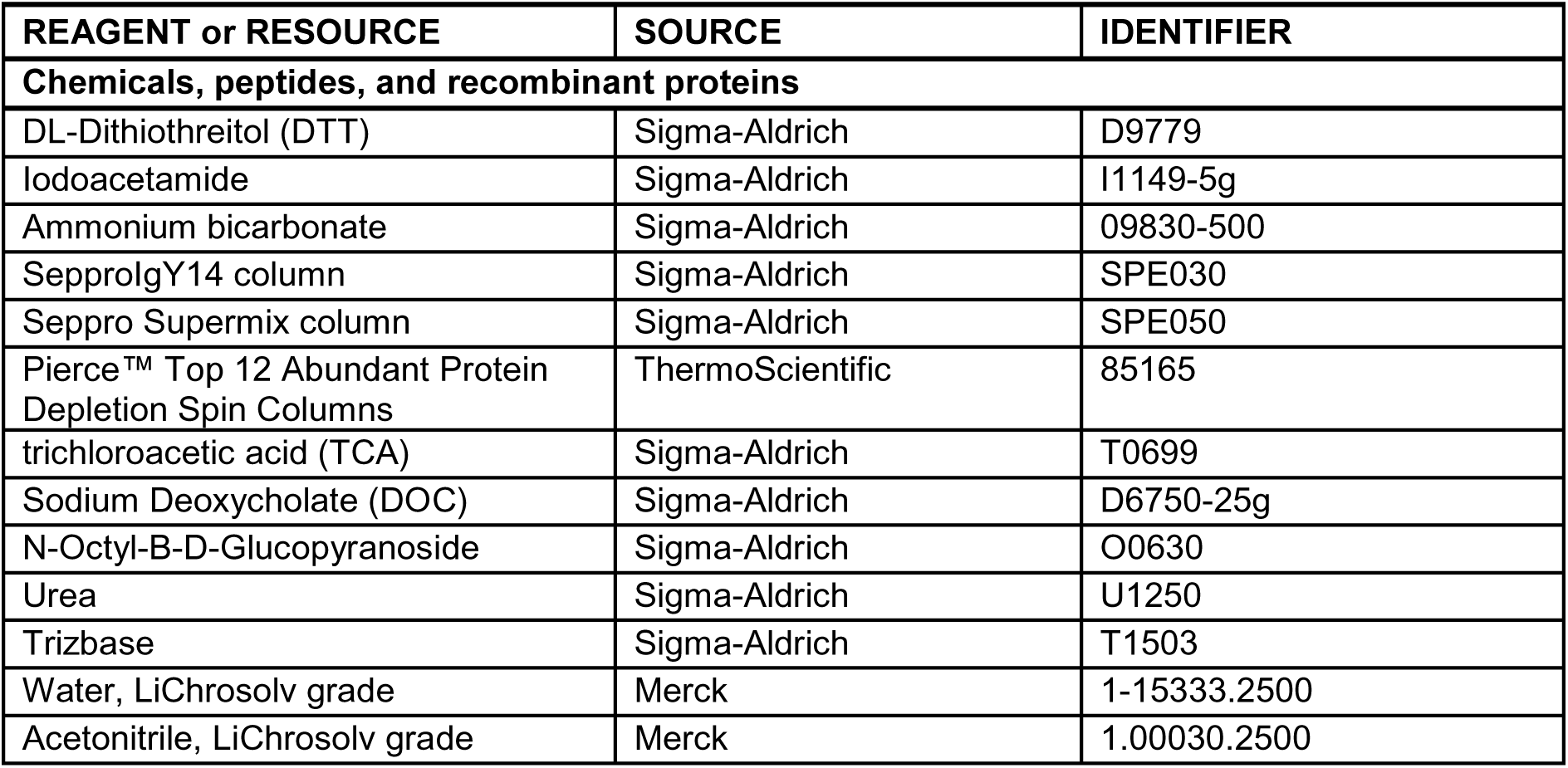
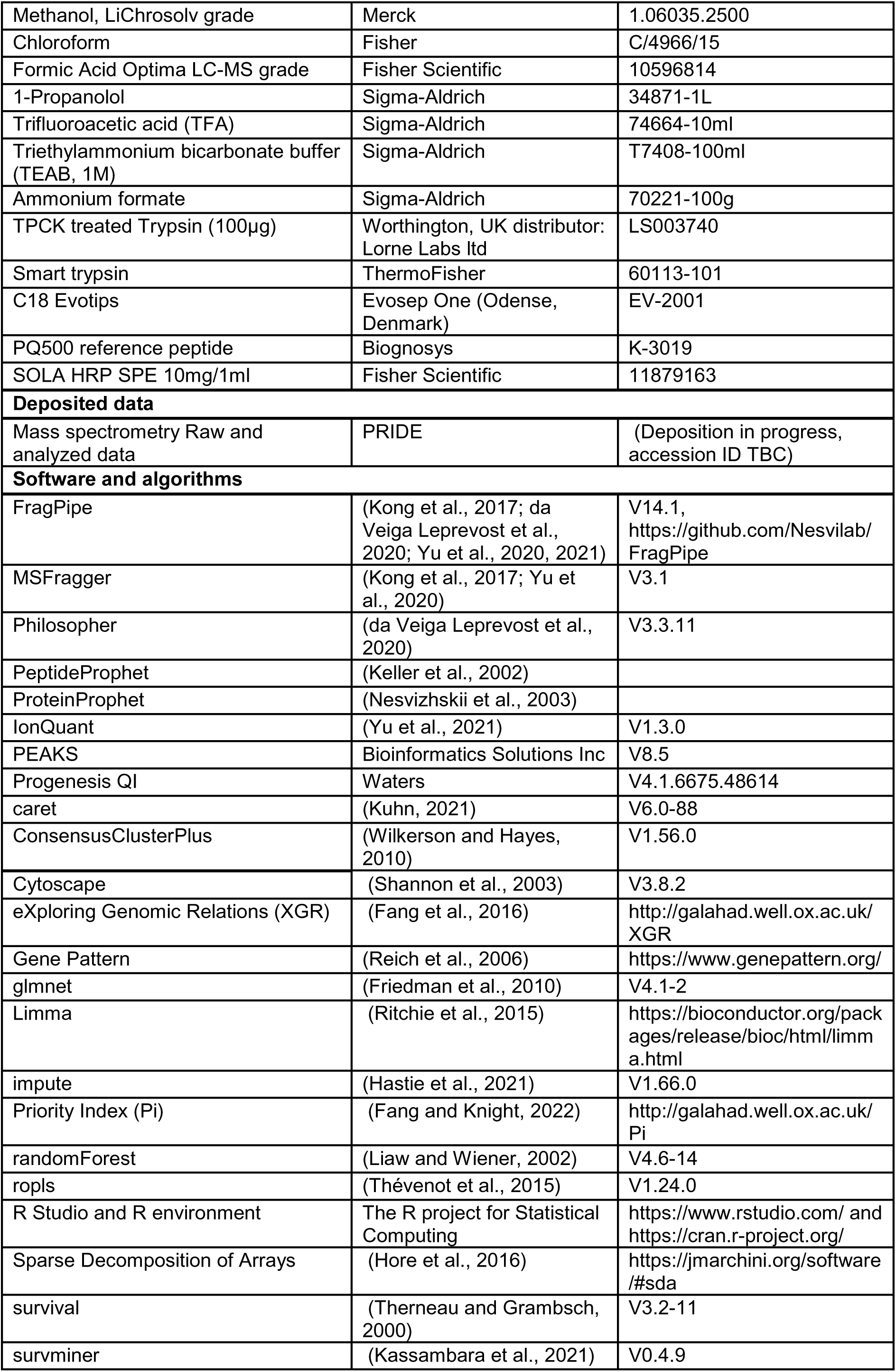
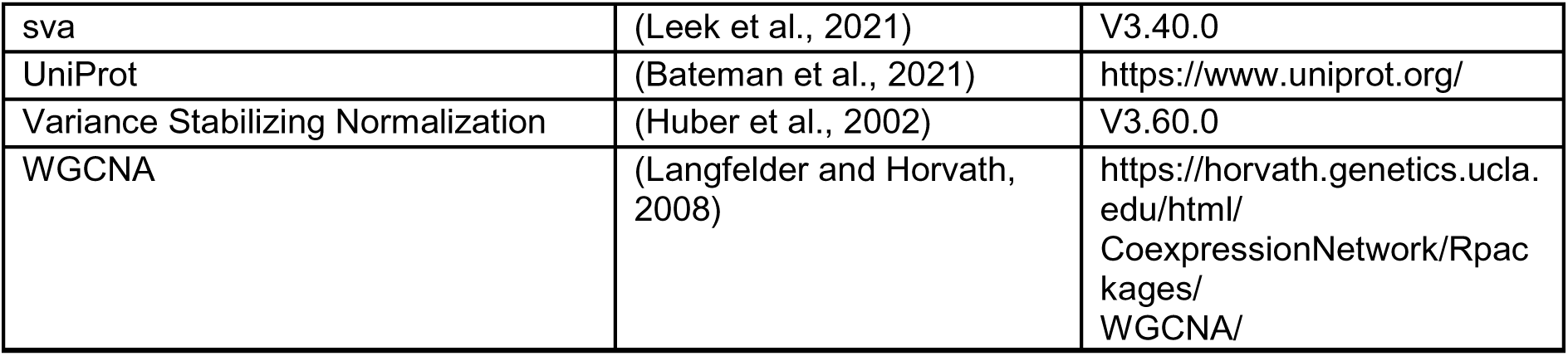

## Supplementary Tables

**Table S1. Related to Figure 1. Clinical characteristics of cohorts included. Attached as a separate Excel file.**

**Table S2. Related to Figure 3.**
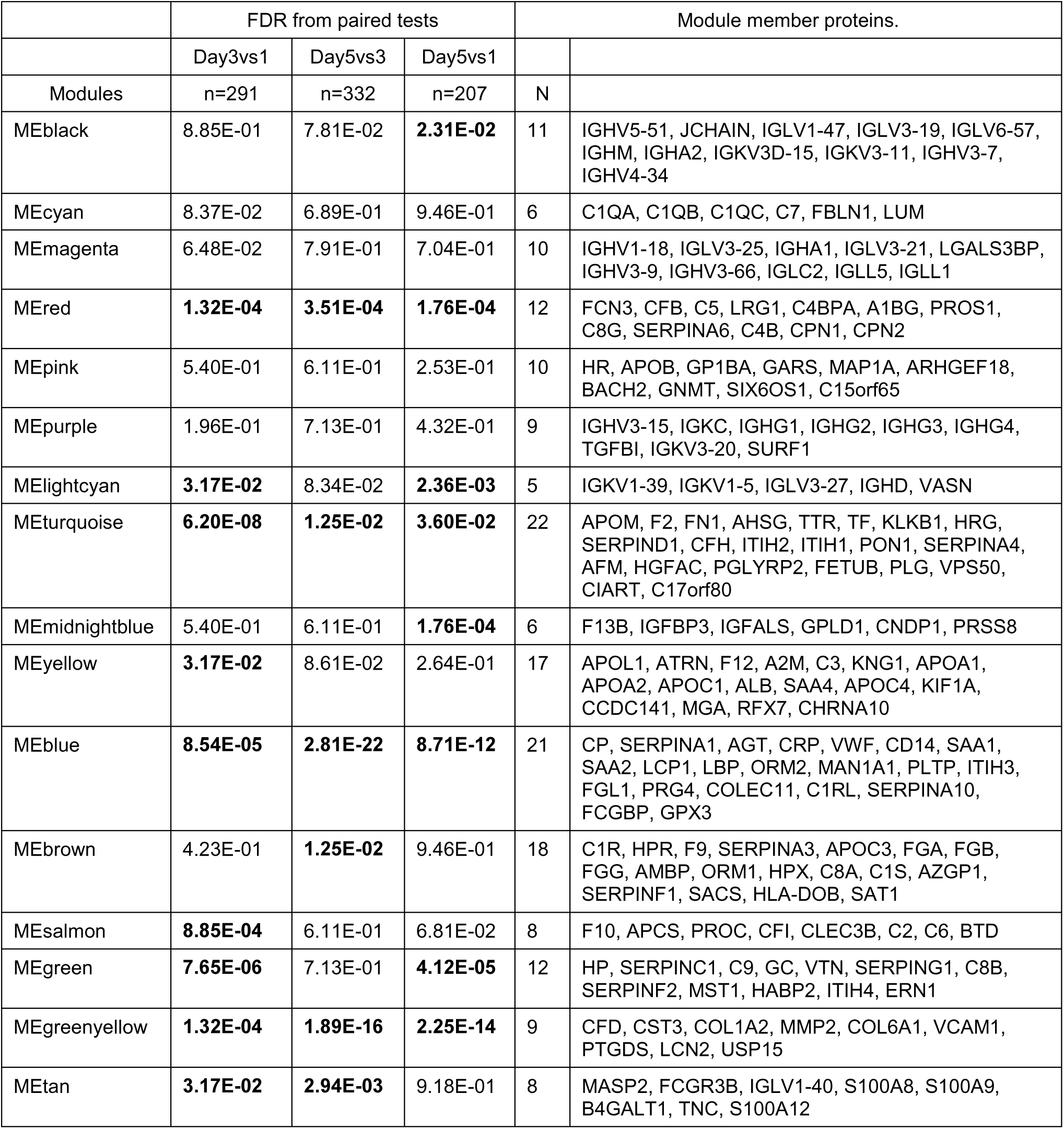
Time-series analysis of protein co-expression modules in Sepsis_ICU. Paired tests of module eigengene (ME) levels between the three time intervals among the first 5 days following ICU admission of sepsis. Numbers of patients with sample pairs available for each interval are showed as n. Multiple testing was corrected for within each interval. Significant differences (FDR<0.05) are indicated in bold.

**Table S3. Related to Figure 4.** Comparison of clinical variables between sepsis discovery cohort clusters. Only variables with a significant difference between the clusters are shown. Abbreviations are used as in Table S2. K-W = Kruskal-Wallis test. Dunn’s = Dunn’s post-hoc test. “Dunn’s FDR” shows the p values corrected within the three pairs tested for each variable, in the order of “Dunn’s test pairs”.

|  |  | No. / total patients with information (%) |  |  |
| --- | --- | --- | --- | --- |
| Clinical variable | $\chi^2$ FDR | SPC1 | SPC2 | SPC3 |
| Diagnosis - FP | 5.00E-12 | 99/169 (59) | 51/180 (28) | 117/434 (27) |
| Timepoint | 0.0024 |  |  |  |
| - Day1 |  | 109/170 (64) | 87/183 (48) | 210/435 (48) |
| - Day5 |  | 10/170 (6) | 31/183 (17) | 77/435 (18) |
| Shock | 1.40E-07 | 130/168 (77) | 87/179 (49) | 231/426 (54) |
| Mechanical ventilation/CPAP | 0.015 | 126/169 (75) | 114/181 (63) | 260/431 (60) |
| Acute renal failure | 3.40E-08 | 64/169 (38) | 30/181 (17) | 68/431 (16) |

|  |  |  |  | Median (IQR) |  |  | N(patients) with information |  |  |
| --- | --- | --- | --- | --- | --- | --- | --- | --- | --- |
| Clinical variable | K-W FDR | Dunn's FDR | Dunn's test pairs | SPC1 | SPC2 | SPC3 | SPC1 | SPC2 | SPC3 |
| Age | 0.014 | 0.012, 0.020 | SPC1vs3, SPC2vs3 | 67 (56-77) | 67 (53-77) | 63 (51-74) | 170 | 183 | 435 |
| Estimated day from onset (FP) | 0.062 | 0.049, 0.028 | SPC1vs2, SPC1vs3 | 2 (1-3) | 3.0 (1.2-4.0) | 3 (1-5) | 99 | 50 | 116 |
| Systolic blood pressure (low) | 9.6E-06 | 5.1e-05, 1.4e-06 | SPC1vs2, SPC1vs3 | 94 ( 82-102) | 101 ( 90-110) | 100 ( 89-113) | 169 | 180 | 430 |
| Mean arterial pressure (low) | 4.6E-06 | 2.7e-05, 6.6e-07 | SPC1vs2, SPC1vs3 | 62 (56-70) | 66 (61-75) | 67 (60-75) | 168 | 180 | 426 |
| Heart rate (high) | 0.014 | 0.0275, 0.0035 | SPC1vs2, SPC1vs3 | 115 (100-129) | 108 ( 95-123) | 107 ( 95-124) | 169 | 180 | 430 |
| Inotropic support (days) | 2.3E-06 | 1.6e-06, 1.5e-06 | SPC1vs2, SPC1vs3 | 3 (1-6) | 1 (0-3) | 1 (0-4) | 169 | 183 | 434 |
| Arterial pH | 0.0001 | 4.5e-05, 1.7e-04 | SPC1vs2, SPC1vs3 | 7.3 (7.2-7.4) | 7.4 (7.3-7.4) | 7.4 (7.3-7.4) | 108 | 86 | 209 |
| Respiratory rate | 0.000026 | 4.5e-06, 1.0e-04, 4.1e-02 | SPC1vs2, SPC1vs3, SPC2vs3 | 23 (18-29) | 27 (21-35) | 26 (20-33) | 168 | 180 | 430 |
| Partial pressure of oxygen | 0.00049 | 5.1e-03, 6.3e-05 | SPC1vs2, SPC1vs3 | 9.4 ( 8.4-10.8) | 8.9 ( 7.9-10.2) | 8.8 ( 7.8-10.1) | 166 | 168 | 403 |
| Lactate | 3.1E-13 | 5.5e-09, 4.4e-15 | SPC1vs2, SPC1vs3 | 2.9 (1.8-4.3) | 1.6 (1.2-2.3) | 1.6 (1.2-2.2) | 111 | 115 | 265 |
| Bicarbonate | 1.6E-12 | 3.1e-09, 1.2e-13 | SPC1vs2, SPC1vs3 | 21 (18-24) | 25 (21-28) | 25 (21-28) | 168 | 170 | 409 |
| Urea (high) | 1.2E-11 | 2.0e-08, 9.7e-13 | SPC1vs2, SPC1vs3 | 12.4 ( 8.6-18.0) | 8.0 ( 5.4-14.0) | 8.0 ( 4.9-12.0) | 168 | 180 | 427 |
| Urine volume | 7.4E-16 | 3.1e-13, 9.1e-17 | SPC1vs2, SPC1vs3 | 953 ( 318-1595) | 1910 (1100-2710) | 1730 (1142-2642) | 169 | 177 | 427 |
| Creatinine (high) | 3.4E-13 | 4.1e-09, 7.5e-15 | SPC1vs2, SPC1vs3 | 123 ( 89-200) | 78 ( 57-126) | 77 ( 56-118) | 169 | 180 | 431 |
| Creatinine (low) | 1.3E-11 | 1.4e-08, 1.4e-12 | SPC1vs2, SPC1vs3 | 108 ( 80-175) | 72 ( 53-109) | 72 ( 53-112) | 169 | 180 | 431 |
| Renal support (days) | 2.8E-09 | 1.6e-09, 1.9e-08, 4.9e-02 | SPC1vs2, SPC1vs3, SPC2vs3 | 0 (0-2) | 0 (0-0) | 0 (0-0) | 169 | 183 | 434 |
| Prothrombin time | 0.0012 | 0.03199, 0.00017 | SPC1vs2, SPC1vs3 | 16 (14-20) | 15 (12-17) | 14 (12-16) | 68 | 56 | 141 |
| The international normalized ratio | 0.000026 | 1.1e-04, 3.3e-06 | SPC1vs2, SPC1vs3 | 1.3 (1.2-1.7) | 1.2 (1.1-1.3) | 1.2 (1.1-1.3) | 67 | 88 | 201 |
| Bilirubin (high) | 0.0054 | 0.0033, 0.0020 | SPC1vs2, SPC1vs3 | 13 ( 8-22) | 10 ( 6-16) | 10 ( 6-17) | 163 | 174 | 416 |
| Temperature (low) | 0.0025 | 0.00222, 0.00075 | SPC1vs2, SPC1vs3 | 36 (36-36) | 36 (36-37) | 36 (36-37) | 169 | 180 | 430 |
| Platelets (low) | 0.00034 | 3.7e-03, 4.1e-05 | SPC1vs2, SPC1vs3 | 172 (106-265) | 205 (146-280) | 215 (156-298) | 169 | 179 | 431 |
| White cell count (low) | 0.0038 | 4e-02, 7e-04 | SPC1vs2, SPC1vs3 | 9.7 ( 5.7-15.0) | 11.4 ( 7.5-15.2) | 12 ( 8-16) | 169 | 180 | 431 |
| Hematocrit | 3.4E-08 | 1.8e-06, 9.4e-09 | SPC1vs2, SPC1vs3 | 31 (28-35) | 35 (32-40) | 36 (31-41) | 92 | 71 | 175 |
| Lymphocytes (raw) | 0.00082 | 0.03499, 0.00014, 0.04981 | SPC1vs2, SPC1vs3, SPC2vs3 | 0.70 (0.48-1.00) | 0.8 (0.5-1.2) | 0.90 (0.58-1.39) | 160 | 177 | 421 |
| Monocytes (raw) | 0.00012 | 2.3e-02, 1.6e-05, 2.7e-02 | SPC1vs2, SPC1vs3, SPC2vs3 | 0.4 (0.2-0.8) | 0.50 (0.33-0.80) | 0.64 (0.36-0.98) | 160 | 177 | 422 |
| Monocytes (proportion) | 0.0097 | 0.032, 0.002 | SPC1vs2, SPC1vs3 | 0.043 (0.023-0.069) | 0.048 (0.030-0.078) | 0.052 (0.032-0.079) | 160 | 177 | 421 |
| APACHE | 0.000029 | 6.8e-06, 2.4e-04, 4.0e-02 | SPC1vs2, SPC1vs3, SPC2vs3 | 16 (12-22) | 12 (10-15) | 14 (10-17) | 107 | 84 | 185 |
| SOFA-Total | 1.2E-11 | 2.3e-10, 1.2e-11 | SPC1vs2, SPC1vs3 | 7 ( 5-10) | 5 (3-7) | 5 (3-7) | 163 | 164 | 391 |
| SOFA-CVS | 1.2E-11 | 6.8e-10, 5.0e-12 | SPC1vs2, SPC1vs3 | 4 (1-4) | 1 (0-3) | 1 (0-3) | 169 | 181 | 431 |
| SOFA-Haem | 0.00002 | 2.0e-04, 2.1e-06 | SPC1vs2, SPC1vs3 | 0 (0-1) | 0 (0-1) | 0 (0-0) | 169 | 179 | 431 |
| SOFA-Hep | 0.0015 | 0.00089, 0.00055 | SPC1vs2, SPC1vs3 | 0 (0-1) | 0 (0-0) | 0 (0-0) | 164 | 174 | 416 |
| SOFA-Renal | 1.2E-12 | 5.5e-10, 1.1e-13 | SPC1vs2, SPC1vs3 | 1 (0-4) | 0 (0-1) | 0 (0-1) | 169 | 181 | 431 |
| SOFA-Resp | 0.0031 | 0.0013, 0.0019 | SPC1vs2, SPC1vs3 | 2 (2-2) | 2 (2-2) | 2 (2-2) | 168 | 170 | 404 |

**Table S4. Related to Figure 5.** Patient numbers for proteomic cluster movement. Tables show number of patients who moved between specified proteomic clusters at specified sampling timepoints, in the sepsis discovery and validation cohorts. Patients with samples available at both specified timepoints are counted in each table.

| From Day1 to Day3 |  |  |  |
| --- | --- | --- | --- |
|  | To SPC1 | To SPC2 | To SPC3 |
| From SPC1 | 43 | 19 | 24 |
| From SPC2 | 7 | 28 | 26 |
| From SPC3 | 10 | 26 | 108 |
| From Day3 to Day5 |  |  |  |
|  | To SPC1 | To SPC2 | To SPC3 |
| From SPC1 | 50 | 15 | 25 |
| From SPC2 | 2 | 36 | 35 |
| From SPC3 | 5 | 36 | 128 |
| From Day1 to Day5 |  |  |  |
|  | To SPC1 | To SPC2 | To SPC3 |
| From SPC1 | 21 | 17 | 29 |
| From SPC2 | 2 | 20 | 18 |
| From SPC3 | 6 | 16 | 78 |

**Table S5. Related to Figure 5.** Evidence for each entry in the summary graph of molecular characteristics of the clusters. Pathway enrichment refers to enrichment of proteins with either significantly higher or lower abundance in each contrast, using GOBP or Reactome annotations. Levels indicated were verified by visual examinations of the distributions across the clusters for the overlapping differentially abundant proteins. ^1^This is a general term including phagocytosis, Fc-γ/ɛ receptor signaling, leukocyte migration, defense response to bacterium, and complement activation of classical pathway. The overlapping proteins annotated for these pathways as well as for “B cell signaling” were mainly constituted of immunoglobulins including the constant and variable regions. Abbreviations: disc – discovery cohort; vali – validation cohort; HV – healthy volunteers; DA – differentially abundant.

| Term listed | Cluster | Level | Evidence from |  |  |
| --- | --- | --- | --- | --- | --- |
|  |  |  | Dataset | Contrast | Approach |
| USP15, COL1A2 | SPC1 | up | disc, vali | SPC1vs3, SPC1vs2 | Top DA proteins |
| MAP1A, GNMT | SPC2 | down | disc, vali | SPC2vs3, SPC1vs2 | Top DA proteins |
| TSPAN11 | SPC2 | up | disc, vali | SPC2vs3, SPC1vs2 | Top DA proteins |
| Cytokines and chemokines | SPC1 | up | Luminex | SPC1vs3 | Individual DA proteins |
|  | SPC3 | down | Luminex | SPC1vs3 | Individual DA proteins |
| Regulation of immune response<sup>1</sup> | SPC1 | up | disc, vali, QE-HF | SPC1vs2, SPC1vs3 | Pathway enrichment |
| Immunoglobulins | SPC1 | up | disc, vali | SPC1vs2 | Individual DA proteins |
|  | SPC2 | down | disc, vali | SPC1vs2 | Individual DA proteins |
| B cell signaling | SPC1 | up | disc, vali | SPC1vs2 | Pathway enrichment |
|  |  |  | disc | SPC1vsHV | Pathway enrichment |
|  |  |  | Luminex | SPC1vs3 | Individual protein functions |
|  | SPC2 | down | disc, vali | SPC1vs2 | Pathway enrichment |
| Interleukin signaling | SPC1 | up | disc, vali | SPC1vs2, SPC1vs3 | Pathway enrichment |
|  |  |  | QE-HF | SPC1vs3 | Pathway enrichment |
|  | SPC3 | down | QE-HF | SPC1vs3 | Pathway enrichment |
| ECM organization | SPC1 | up | disc, vali | SPC1vs2, SPC1vs3 | Pathway enrichment |
|  |  |  | QE-HF | SPC1vs3 | Pathway enrichment |
|  | SPC3 | down | QE-HF | SPC1vs3 | Pathway enrichment |
| Collagen fibril organization | SPC1 | up | QE-HF | SPC1vs3 | Pathway enrichment |
|  | SPC3 | down | QE-HF | SPC1vs3 | Pathway enrichment |
| Cell division | SPC1 | up | QE-HF | SPC1vs3 | Pathway enrichment |
|  | SPC3 | down | QE-HF | SPC1vs3 | Pathway enrichment |
| Lipoprotein metabolic process | SPC1 | down | disc, vali | SPC1vs2 | Pathway enrichment |
|  |  |  | QE-HF | SPC1vs3 | Pathway enrichment |
|  | SPC2 | up | disc, vali | SPC1vs2 | Pathway enrichment |
|  | SPC3 | up | QE-HF | SPC1vs3 | Pathway enrichment |
| Transport of small molecules | SPC1 | down | disc | SPC1vs2 | Pathway enrichment |
|  |  |  | QE-HF | SPC1vs3 | Pathway enrichment |
|  | SPC2 | up | disc | SPC1vs2 | Pathway enrichment |
|  | SPC3 | up | QE-HF | SPC1vs3 | Pathway enrichment |

**Table S6. Related to Figure 6.** Matrix decomposition for integrating plasma proteome and leukocyte transcriptome in sepsis. Attached as a separate Excel file.

**Table S7. Related to Figure 7.**
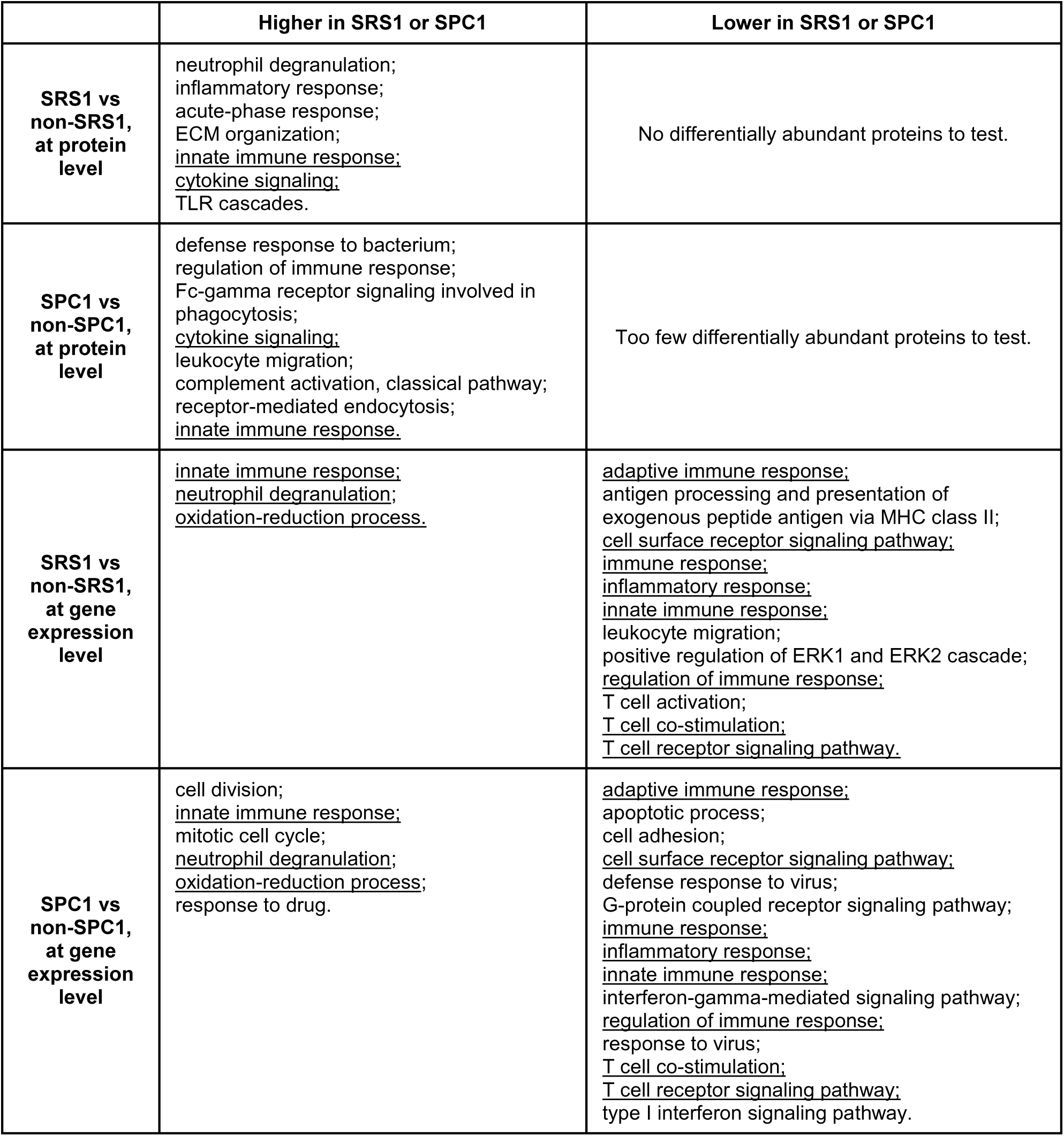
Comparison of enriched pathways in SRS1 or SPC1 contrasts. Significantly enriched terms (FDR<0.05) are listed in the table. Terms shared between the SRS1 and SPC1 contrasts are underlined. At protein level, top 10% proteins by significance were tested for enrichment against the total 269 proteins as background, using both GOBP and Reactome annotations. A minimum of 5 overlaps between input data and genes in the terms was required for the term to be tested. At gene expression level, top 2% genes by significance were tested for enrichment against the total genes as background, using only GOBP annotations. A minimum of 10 overlaps between input data and genes in the terms was required for the term to be tested. Redundant terms have been removed by the function xEnrichConciser from package XGR. Only terms identified in both microarray and RNAseq data are listed.

## Notes

### Author Declarations

Ethical approval for this work was given by the following UK Research Ethics Committees: Scotland A Research Ethics Committee (05/MRE00/38), Berkshire Research Ethics Committee (08/H0505/78), South Central - Oxford A Research Ethics Committee (12/SC/0014), South Central - Oxford C Research Ethics Committee (18/SC/0588), London - Stanmore Research Ethics Committee (16/LO/0635), East Midlands - Nottingham 2 Research Ethics Committee (14/EM/1223), London - Harrow Research Ethics Committee (14/LO/2004), London - Camden and Kings Cross Research Ethics Committee (15/LO/0933), and North London Research Ethics Committee (10/H0709/77).

## References

Aebersold, R., and Mann, M. (2003). Mass spectrometry-based proteomics. Nature 422, 198–207.

Angus, D.C., Marrie, T.J., Scott Obrosky, D., Clermont, G., Dremsizov, T.T., Coley, C., Fine, M.J., Singer, D.E., and Kapoor, W.N. (2002). Severe community-acquired pneumonia: Use of intensive care services and evaluation of American and British Thoracic Society diagnostic criteria. Am. J. Respir. Crit. Care Med. 166, 717–723.

Antcliffe, D., Jiménez, B., Veselkov, K., Holmes, E., and Gordon, A.C. (2017). Metabolic Profiling in Patients with Pneumonia on Intensive Care. EBioMedicine 18, 244–253.

Antcliffe, D.B., Wolfer, A.M., O’Dea, K.P., Takata, M., Holmes, E., and Gordon, A.C. (2018). Profiling inflammatory markers in patients with pneumonia on intensive care. Sci. Rep. 8, 1–9.

Antcliffe, D.B., Burnham, K.L., Al-Beidh, F., Santhakumaran, S., Brett, S.J., Hinds, C.J., Ashby, D., Knight, J.C., and Gordon, A.C. (2019). Transcriptomic signatures in sepsis and a differential response to steroids from the VaNISH randomized trial. Am. J. Respir. Crit. Care Med. 199, 980– 986.

Antcliffe, D.B., Mi, Y., Santhakumaran, S., Burnham, K.L., Prevost, T., Ward, J., Marshall, T., Bradley, C., Al-Beidh, F., Hutton, P., et al. (2022). Inflammatory sub-phenotypes in sepsis: relationship to outcomes, treatment effect and transcriptomic sub-phenotypes. MedRxiv 2022.07.12.22277463.

Bache, N., Hoerning, O., Falkenby, L., and Vorm, O. EVOSEP ONE: A GRADIENT OFF-SET FOCUSING UHPLC INSTRUMENT FOR ROBUST AND HIGH THROUGHPUT PROTEOMICS.

Baghela, A., Pena, O.M., Lee, A.H., Baquir, B., Falsafi, R., An, A., Farmer, S.W., Hurlburt, A., Mondragon-Cardona, A., Rivera, J.D., et al. (2022). Predicting sepsis severity at first clinical presentation: The role of endotypes and mechanistic signatures. EBioMedicine 75, 103776.

Bateman, A., Martin, M.J., Orchard, S., Magrane, M., Agivetova, R., Ahmad, S., Alpi, E., Bowler-Barnett, E.H., Britto, R., Bursteinas, B., et al. (2021). UniProt: the universal protein knowledgebase in 2021. Nucleic Acids Res. 49, D480–D489.

Beer, L.A., Ky, B., Barnhart, K.T., and Speicher, D.W. (2017). In-Depth, Reproducible Analysis of Human Plasma Using IgY 14 and SuperMix Immunodepletion. Methods Mol. Biol. 1619, 81–101.

Brunson, J.C. (2020). {ggalluvial}: Layered Grammar for Alluvial Plots. J. Open Source Softw. 5, 2017.

Burnham, K.L., Davenport, E.E., Radhakrishnan, J., Humburg, P., Gordon, A.C., Hutton, P., Svoren-Jabalera, E., Garrard, C., Hill, A.V.S., Hinds, C.J., et al. (2017). Shared and Distinct Aspects of the Sepsis Transcriptomic Response to Fecal Peritonitis and Pneumonia. Am. J. Respir. Crit. Care Med. 196, 328–339.

Cano-Gamez, E., Burnham, K.L., Goh, C., Malick, Z.H., Kwok, A., Smith, D.A., Peters-Sengers, H., Antcliffe, D., Investigators, Ga., McKechnie, S., et al. (2022). An immune dysfunction score for stratification of patients with acute infection based on whole blood gene expression. MedRxiv 2022.03.17.22272427.

Chawade, A., Alexandersson, E., and Levander, F. (2014). Normalyzer: a tool for rapid evaluation of normalization methods for omics data sets. J. Proteome Res. 13, 3114–3120.

Chidambaram, V., Shanmugavel Geetha, H., Kumar, A., Majella, M.G., Sivakumar, R.K., Voruganti, D., Mehta, J.L., and Karakousis, P.C. (2022). Association of Lipid Levels With COVID-19 Infection, Disease Severity and Mortality: A Systematic Review and Meta-Analysis. Front. Cardiovasc. Med. 9.

De Coux, A., Tian, Y., Deleon-Pennell, K.Y., Nguyen, N.T., De Castro Brás, L.E., Flynn, E.R., Cannon, P.L., Griswold, M.E., Jin, Y.F., Puskarich, M.A., et al. (2015). Plasma glycoproteomics reveals sepsis outcomes linked to distinct proteins in common pathways. Crit. Care Med. 43, 2049– 2058.

COvid-19 Multi-omics Blood ATlas (COMBAT) Consortium (2022). A blood atlas of COVID-19 defines hallmarks of disease severity and specificity. Cell 185, 916–938.e58.

Davenport, E.E., Burnham, K.L., Radhakrishnan, J., Humburg, P., Hutton, P., Mills, T.C., Rautanen, A., Gordon, A.C., Garrard, C., Hill, A.V.S., et al. (2016). Genomic landscape of the individual host response and outcomes in sepsis: A prospective cohort study. Lancet Respir. Med. 4, 259–271.

Fang, H., and Knight, J.C. (2022). Priority index: database of genetic targets in immune-mediated disease. Nucleic Acids Res. 50, D1358–D1367.

Fang, H., Knezevic, B., Burnham, K.L., and Knight, J.C. (2016). XGR software for enhanced interpretation of genomic summary data, illustrated by application to immunological traits. Genome Med. 8, 129.

Fjell, C.D., Thair, S., Hsu, J.L., Walley, K.R., Russell, J.A., and Boyd, J. (2013). Cytokines and signaling molecules predict clinical outcomes in sepsis. PLoS One 8.

Friedman, J., Hastie, T., and Tibshirani, R. (2010). Regularization Paths for Generalized Linear Models via Coordinate Descent. J. Stat. Softw. 33, 1.

Geyer, P.E., Voytik, E., Treit, P. V, Doll, S., Kleinhempel, A., Niu, L., Müller, J.B., Buchholtz, M., Bader, J.M., Teupser, D., et al. (2019). Plasma Proteome Profiling to detect and avoid sample-related biases in biomarker studies. EMBO Mol. Med. 11.

Gordon, A.C., Mason, A.J., Thirunavukkarasu, N., Perkins, G.D., Cecconi, M., Cepkova, M., Pogson, D.G., Aya, H.D., Anjum, A., Frazier, G.J., et al. (2016). Effect of Early Vasopressin vs Norepinephrine on Kidney Failure in Patients With Septic Shock: The VANISH Randomized Clinical Trial. JAMA 316, 509–518.

Graw, S., Chappell, K., Washam, C.L., Gies, A., Bird, J., Robeson, M.S., and Byrum, S.D. (2021). Multi-omics data integration considerations and study design for biological systems and disease. Mol. Omi. 17, 170–185.

Hastie, T., Tibshirani, R., Narasimhan, B., and Chu, G. (2021). impute: impute: Imputation for microarray data.

Hore, V., Viñuela, A., Buil, A., Knight, J., McCarthy, M.I., Small, K., and Marchini, J. (2016). Tensor decomposition for multiple-tissue gene expression experiments. Nat. Genet. 48, 1094–1100.

Huber, W., Von Heydebreck, A., Ultmann, H.S., Poustka, A., and Vingron, M. (2002). Variance stabilization applied to microarray data calibration and to the quantification of differential expression.

Karp, N.A., Huber, W., Sadowski, P.G., Charles, P.D., Hester, S. V, and Lilley, K.S. (2010). Addressing accuracy and precision issues in iTRAQ quantitation. Mol. Cell. Proteomics 9, 1885– 1897.

Karpe, F., Vasan, S.K., Humphreys, S.M., Miller, J., Cheeseman, J., Dennis, A.L., and Neville, M.J. (2018). Cohort Profile: The Oxford Biobank Why was the cohort set up? Int. J. Epidemiol.

Kassambara, A., Kosinski, M., and Biecek, P. (2021). survminer: Drawing Survival Curves using “ggplot2.”

Keller, A., Nesvizhskii, A.I., Kolker, E., and Aebersold, R. (2002). Empirical statistical model to estimate the accuracy of peptide identifications made by MS/MS and database search. Anal. Chem. 74, 5383–5392.

Keshishian, H., Burgess, M.W., Specht, H., Wallace, L., Clauser, K.R., Gillette, M.A., and Carr, S.A. (2017). Quantitative, multiplexed workflow for deep analysis of human blood plasma and biomarker discovery by mass spectrometry. Nat. Protoc. 12, 1683–1701.

Kiehntopf, M., Schmerler, D., Brunkhorst, F.M., Winkler, R., Ludewig, K., Osterloh, D., Bloos, F., Reinhart, K., and Deufel, T. (2011). Mass Spectometry-Based Protein Patterns in the Diagnosis of Sepsis/Systemic Inflammatory Response Syndrome. Shock 36, 560–569.

Kivelä, M., Arenas, A., Barthelemy, M., Gleeson, J.P., Moreno, Y., and Porter, M.A. (2014). Multilayer networks. J. Complex Networks 2, 203–271.

Kong, A.T., Leprevost, F. V., Avtonomov, D.M., Mellacheruvu, D., and Nesvizhskii, A.I. (2017). MSFragger: ultrafast and comprehensive peptide identification in mass spectrometry–based proteomics. Nat. Methods 2017 145 14, 513–520.

Kuhn, M. (2021). caret: Classification and Regression Training.

Kuroda, K., Wake, H., Mori, S., Hinotsu, S., Nishibori, M., and Morimatsu, H. (2018). Decrease in Histidine-Rich Glycoprotein as a Novel Biomarker to Predict Sepsis Among Systemic Inflammatory Response Syndrome. Crit. Care Med. 46, 570–576.

Kwok, A.J., Allcock, A., Ferreira, R.C., Smee, M., Cano-Gamez, E., Burnham, K.L., Zurke, Y.X., research, O. acute medicine/ED, McKechnie, S., Monaco, C., et al. (2022). Identification of deleterious neutrophil states and altered granulopoiesis in sepsis. MedRxiv 2022.03.22.22272723.

Langfelder, P., and Horvath, S. (2008). WGCNA: An R package for weighted correlation network analysis. BMC Bioinformatics 9, 1–13.

Langley, R.J., Tsalik, E.L., Van Velkinburgh, J.C., Glickman, S.W., Rice, B.J., Wang, C., Chen, B., Carin, L., Suarez, A., Mohney, R.P., et al. (2013). Sepsis: An integrated clinico-metabolomic model improves prediction of death in sepsis. Sci. Transl. Med. 5.

Leek, J.T., Johnson, W.E., Parker, H.S., Fertig, E.J., Jaffe, A.E., Zhang, Y., Storey, J.D., and Torres, L.C. (2021). sva: Surrogate Variable Analysis.

Leite, G.G.F., Ferreira, B.L., Tashima, A.K., Nishiduka, E.S., Cunha-neto, E., Karina, M., Brunialti, C., Scicluna, B.P., and Salomão, R. (2021). Combined Transcriptome and Proteome Leukocyte ’ s Pro fi ling Reveals Up-Regulated Module of Genes / Proteins Related to Low Density Neutrophils and Impaired Transcription and Translation Processes in Clinical Sepsis. 12, 1–14.

Levy, M.M., Fink, M.P., Marshall, J.C., Abraham, E., Angus, D., Cook, D., Cohen, J., Opal, S.M., Vincent, J.L., and Ramsay, G. (2003). 2001 SCCM/ESICM/ACCP/ATS/SIS International Sepsis Definitions Conference. Crit. Care Med. 31, 1250–1256.

Li, Z., Yin, M., Zhang, H., Ni, W., Pierce, R.W., Zhou, H.J., and Min, W. (2020). BMX Represses Thrombin-PAR1-Mediated Endothelial Permeability and Vascular Leakage During Early Sepsis. Circ. Res. 126, 471–485.

Liaw, A., and Wiener, M. (2002). Classification and Regression by randomForest. R News 2, 18–22.

Lorente, L., Martín, M.M., Labarta, L., Díaz, C., Solé-Violán, J., Blanquer, J., Orbe, J., Rodríguez, J.A., Jiménez, A., Borreguero-León, J.M., et al. (2009). Matrix metalloproteinase-9, -10, and tissue inhibitor of matrix metalloproteinases-1 blood levels as biomarkers of severity and mortality in sepsis. Crit. Care 13.

Marshall, J.C. (2014). Why have clinical trials in sepsis failed? Trends Mol. Med. 20, 195–203.

Maslove, D.M., Tang, B., Shankar-Hari, M., Lawler, P.R., Angus, D.C., Baillie, J.K., Baron, R.M., Bauer, M., Buchman, T.G., Calfee, C.S., et al. (2022). Redefining critical illness. Nat. Med. 2022 286 28, 1141–1148.

Matsushita, M. (2010). Ficolins: complement-activating lectins involved in innate immunity. J. Innate Immun. 2, 24–32.

May, S.M., Reyes, A., Martir, G., Reynolds, J., Paredes, L.G., Karmali, S., Stephens, R.C.M., Brealey, D., and Ackland, G.L. (2019). Acquired loss of cardiac vagal activity is associated with myocardial injury in patients undergoing noncardiac surgery: prospective observational mechanistic cohort study. Br. J. Anaesth. 123, 758–767.

May, S.M., Abbott, T.E.F., Del Arroyo, A.G., Reyes, A., Martir, G., Stephens, R.C.M., Brealey, D., Cuthbertson, B.H., Wijeysundera, D.N., Pearse, R.M., et al. (2020). MicroRNA signatures of perioperative myocardial injury after elective noncardiac surgery: a prospective observational mechanistic cohort study. Br. J. Anaesth. 125, 661.

Melani, R.D., Gerbasi, V.R., Anderson, L.C., Sikora, J.W., Toby, T.K., Hutton, J.E., Butcher, D.S., Negrão, F., Seckler, H.S., Srzentić, K., et al. (2022). The Blood Proteoform Atlas: A reference map of proteoforms in human hematopoietic cells. Science (80-.). 375, 411–418.

Miao, H., Chen, S., and Ding, R. (2021). Evaluation of the Molecular Mechanisms of Sepsis Using Proteomics. Front. Immunol. 12.

Morris, J.H., Apeltsin, L., Newman, A.M., Baumbach, J., Wittkop, T., Su, G., Bader, G.D., and Ferrin, T.E. (2011). ClusterMaker: A multi-algorithm clustering plugin for Cytoscape. BMC Bioinformatics 12, 1–14.

Nesvizhskii, A.I., Keller, A., Kolker, E., and Aebersold, R. (2003). A statistical model for identifying proteins by tandem mass spectrometry. Anal. Chem. 75, 4646–4658.

O’Callaghan, D.J.P., O’Dea, K.P., Scott, A.J., Takata, M., and Gordon, A.C. (2015). Monocyte tumor necrosis factor-α-converting enzyme catalytic activity and substrate shedding in sepsis and noninfectious systemic inflammation. Crit. Care Med. 43, 1375–1385.

Patel, P.B., Brett, S.J., O’Callaghan, D., Anjum, A., Cross, M., Warwick, J., and Gordon, A.C. (2020). Methylnaltrexone for the treatment of opioid-induced constipation and gastrointestinal stasis in intensive care patients. Results from the MOTION trial. Intensive Care Med. 46, 747–755.

Pimienta, G., Heithoff, D.M., Rosa-Campos, A., Tran, M., Esko, J.D., Mahan, M.J., Marth, J.D., and Smith, J.W. (2019). Plasma Proteome Signature of Sepsis: a Functionally Connected Protein Network. Proteomics 19, 1800389.

van der Poll, T., Shankar-Hari, M., and Wiersinga, W.J. (2021). The immunology of sepsis. Immunity 54, 2450–2464.

Prianichnikov, N., Koch, H., Koch, S., Lubeck, M., Heilig, R., Brehmer, S., Fischer, R., and Cox, J. (2020). MaxQuant Software for Ion Mobility Enhanced Shotgun Proteomics. Mol. Cell. Proteomics 19, 1058–1069.

R Core Team (2015). R: a language and environment for statistical computing (R Foundation for Statistical Computing).

Rajczewski, A.T., Jagtap, P.D., and Griffin, T.J. (2022). An overview of technologies for MS-based proteomics-centric multi-omics. Expert Rev. Proteomics 1–17.

Rautanen, A., Mills, T.C., Gordon, A.C., Hutton, P., Steffens, M., Nuamah, R., Chiche, J.D., Parks, T., Chapman, S.J., Davenport, E.E., et al. (2015). Genome-wide association study of survival from sepsis due to pneumonia: An observational cohort study. Lancet Respir. Med. 3, 53–60.

Reich, M., Liefeld, T., Gould, J., Lerner, J., Tamayo, P., and Mesirov, J.P. (2006). GenePattern 2.0. Nat. Genet. 38, 500–501.

Ritchie, M.E., Phipson, B., Wu, D., Hu, Y., Law, C.W., Shi, W., and Smyth, G.K. (2015). limma powers differential expression analyses for RNA-sequencing and microarray studies. Nucleic Acids Res. 43, e47.

Rudd, K.E., Johnson, S.C., Agesa, K.M., Shackelford, K.A., Tsoi, D., Kievlan, D.R., Colombara, D. V., Ikuta, K.S., Kissoon, N., Finfer, S., et al. (2020). Global, regional, and national sepsis incidence and mortality, 1990–2017: analysis for the Global Burden of Disease Study. Lancet 395, 200–211.

Scicluna, B.P., van Vught, L.A., Zwinderman, A.H., Wiewel, M.A., Davenport, E.E., Burnham, K.L., Nürnberg, P., Schultz, M.J., Horn, J., Cremer, O.L., et al. (2017). Classification of patients with sepsis according to blood genomic endotype: a prospective cohort study. Lancet Respir. Med. 5, 816–826.

Seymour, C.W., Kennedy, J.N., Wang, S., Chang, C.C.H., Elliott, C.F., Xu, Z., Berry, S., Clermont, G., Cooper, G., Gomez, H., et al. (2019). Derivation, Validation, and Potential Treatment Implications of Novel Clinical Phenotypes for Sepsis. In JAMA - Journal of the American Medical Association, (American Medical Association), pp. 2003–2017.

Shannon, P., Markiel, A., Ozier, O., Baliga, N.S., Wang, J.T., Ramage, D., Amin, N., Schwikowski, B., and Ideker, T. (2003). Cytoscape: a software environment for integrated models of biomolecular interaction networks. Genome Res. 13, 2498–2504.

Sharma, N.K., Tashima, A.K., Brunialti, M.K.C., Ferreira, E.R., Torquato, R.J.S., Mortara, R.A., Machado, F.R., Assuncao, M., Rigato, O., and Salomao, R. (2017). Proteomic study revealed cellular assembly and lipid metabolism dysregulation in sepsis secondary to community-acquired pneumonia. Sci. Rep. 7, 15606.

Sharma, N.K., Ferreira, B.L., Tashima, A.K., Brunialti, M.K.C., Torquato, R.J.S., Bafi, A., Assuncao, M., Azevedo, L.C.P., and Salomao, R. (2019). Lipid metabolism impairment in patients with sepsis secondary to hospital acquired pneumonia, a proteomic analysis. Clin. Proteomics 16, 1–13.

Shields-Cutler, R.R., Crowley, J.R., Miller, C.D., Stapleton, A.E., Cui, W., and Henderson, J.P. (2016). Human Metabolome-derived Cofactors Are Required for the Antibacterial Activity of Siderocalin in Urine. J. Biol. Chem. 291, 25901–25910.

Singer, M., Deutschman, C.S., Seymour, C., Shankar-Hari, M., Annane, D., Bauer, M., Bellomo, R., Bernard, G.R., Chiche, J.D., Coopersmith, C.M., et al. (2016). The third international consensus definitions for sepsis and septic shock (sepsis-3). JAMA - J. Am. Med. Assoc. 315, 801–810.

Stanski, N.L., and Wong, H.R. (2019). Prognostic and predictive enrichment in sepsis. Nat. Rev. Nephrol.

Sweeney, T.E., Azad, T.D., Donato, M., Haynes, W.A., Perumal, T.M., Henao, R., Bermejo-Martin, J.F., Almansa, R., Tamayo, E., Howrylak, J.A., et al. (2018). Unsupervised analysis of transcriptomics in bacterial sepsis across multiple datasets reveals three robust clusters. Crit. Care Med. 46, 915–925.

Takahashi, Y., Wake, H., Sakaguchi, M., Yoshii, Y., Teshigawara, K., Wang, D., and Nishibori, M. (2021). Histidine-Rich Glycoprotein Stimulates Human Neutrophil Phagocytosis and Prolongs Survival through CLEC1A. J. Immunol. 206, 737–750.

Tanaka, S., Couret, D., Tran-Dinh, A., Duranteau, J., Montravers, P., Schwendeman, A., and Meilhac, O. (2020). High-density lipoproteins during sepsis: from bench to bedside. Crit. Care 24.

Thavarajah, T., Dos Santos, C.C., Slutsky, A.S., Marshall, J.C., Bowden, P., Romaschin, A., and Marshall, J.G. (2020). The plasma peptides of sepsis. Clin. Proteomics 17, 1–18.

Therneau, T.M., and Grambsch, P.M. (2000). Modeling Survival Data: Extending the Cox Model (New York: Springer).

Thévenot, E.A., Roux, A., Xu, Y., Ezan, E., and Junot, C. (2015). Analysis of the Human Adult Urinary Metabolome Variations with Age, Body Mass Index, and Gender by Implementing a Comprehensive Workflow for Univariate and OPLS Statistical Analyses. J. Proteome Res. 14, 3322–3335.

Toledo, A.G., Golden, G., Campos, A.R., Cuello, H., Sorrentino, J., Lewis, N., Varki, N., Nizet, V., Smith, J.W., and Esko, J.D. (2019). Proteomic atlas of organ vasculopathies triggered by Staphylococcus aureus sepsis. Nat. Commun. 10, 1–13.

Tridente, A., Clarke, G.M., Walden, A., McKechnie, S., Hutton, P., Mills, G.H., Gordon, A.C., Holloway, P.A.H., Chiche, J.D., Bion, J., et al. (2014). Patients with faecal peritonitis admitted to European intensive care units: An epidemiological survey of the GenOSept cohort. Intensive Care Med. 40, 202–210.

Välikangas, T., Suomi, T., and Elo, L.L. (2016). A systematic evaluation of normalization methods in quantitative label-free proteomics. Brief. Bioinform. 19, bbw095.

da Veiga Leprevost, F., Haynes, S.E., Avtonomov, D.M., Chang, H.Y., Shanmugam, A.K., Mellacheruvu, D., Kong, A.T., and Nesvizhskii, A.I. (2020). Philosopher: a versatile toolkit for shotgun proteomics data analysis. Nat. Methods 17, 869–870.

Walden, A.P., Clarke, G.M., McKechnie, S., Hutton, P., Gordon, A.C., Rello, J., Chiche, J.D., Stueber, F., Garrard, C.S., and Hinds, C.J. (2014). Patients with community acquired pneumonia admitted to European intensive care units: An epidemiological survey of the GenOSept cohort. Crit. Care 18, 1–9.

Wang, B., Mezlini, A.M., Demir, F., Fiume, M., Tu, Z., Brudno, M., Haibe-Kains, B., and Goldenberg, A. (2014). Similarity network fusion for aggregating data types on a genomic scale. Nat. Methods 2014 113 11, 333–337.

Wilkerson, M.D., and Hayes, D.N. (2010). ConsensusClusterPlus: a class discovery tool with confidence assessments and item tracking. Bioinformatics 26, 1572–1573.

Wong, H.R., Caldwell, J.T., Cvijanovich, N.Z., Weiss, S.L., Fitzgerald, J.C., Bigham, M.T., Jain, P.N., Schwarz, A., Lutfi, R., Nowak, J., et al. (2019). Prospective clinical testing and experimental validation of the Pediatric Sepsis Biomarker Risk Model. Sci. Transl. Med. 11, 9000.

Yu, F., Haynes, S.E., Teo, G.C., Avtonomov, D.M., Polasky, D.A., and Nesvizhskii, A.I. (2020). Fast Quantitative Analysis of timsTOF PASEF Data with MSFragger and IonQuant. Mol. Cell. Proteomics 19, 1575–1585.

Yu, F., Haynes, S.E., and Nesvizhskii, A.I. (2021). IonQuant Enables Accurate and Sensitive Label-Free Quantification With FDR-Controlled Match-Between-Runs. Mol. Cell. Proteomics 20.

Zador, Z., Landry, A., Cusimano, M.D., and Geifman, N. (2019). Multimorbidity states associated with higher mortality rates in organ dysfunction and sepsis: A data-driven analysis in critical care. Crit. Care 23, 1–11.

